# Effects of an early water, sanitation, hygiene, and nutritional intervention on child development at school age: a 7-year follow-up of a cluster-randomized trial in rural Bangladesh

**DOI:** 10.1101/2025.02.25.25321966

**Authors:** Fahmida Tofail, Helen O. Pitchik, Mahfuza Islam, Rizwana Khan, Abul K. Shoab, Fahmida Akter, Shirina Aktar, Tarique M.N. Huda, Mahbubur Rahman, Peter J. Winch, Stephen P. Luby, Lia C.H. Fernald

**Author notes:** Corresponding Author: Helen Pitchik, PhD, MSc, Division of Epidemiology, School of Public Health, University of California, Berkeley, 2121 Berkeley Way West, Room 5302, Berkeley, CA 94720. co-first authors, contributed equally to this work (Fahmida Tofail, Helen O. Pitchik).

## Abstract

**Background:** A previous cluster-randomized controlled trial in Bangladesh found that individual or combined water, handwashing, sanitation, and nutrition interventions during pregnancy and after birth improved developmental outcomes of children at 1 and 2 years of age. We aimed to determine if these intervention effects were sustained at school-age.

**Methods and Findings:** Pregnant women were enrolled between May 2012 and July 2013 and randomized into chlorinated drinking water (W); improved sanitation (S); handwashing with soap (H); combined WSH; nutrition counselling and provision of lipid-based supplements (N); combined WSH+N, or a passive control arm (C) (N=5,551). We followed-up enrolled mothers and children 5 years after intervention completion. Primary outcomes were child cognition, fine motor abilities, behaviour, school achievement, and executive function; secondary outcomes were maternal mental health and the home environment. We conducted intention to treat analyses using generalized linear models to determine unadjusted and adjusted comparisons between each arm and the control, accounting for pair-matching and block level clustering.

Between September 2019 and February 2021, we re-enrolled 3,832 children. Children in the WSH+N, N, and S arms had improved cognitive scores on one or more domains compared to the control arm, with adjusted effect sizes between 0.10 (95%CI: 0.00, 0.20) and 0.15 (0.03, 0.27). Children in all arms except S had improved prosocial behaviour, with effect sizes between 0.21 (0.07, 0.34) and 0.33 (0.17, 0.49). No intervention effects were observed for fine motor, difficult behaviours, executive functioning, or school achievement. Maternal depressive symptoms were improved in the WSH+N, H, and N arms, and the stimulating home environment was improved in all intervention arms. Data collection for this study was interrupted by a 6-month pause at the start of the COVID-19 pandemic.

**Conclusions:** At 7 years of age, we found small, sustained impacts of early water, sanitation, hygiene, and nutrition interventions on child cognitive and social-emotional outcomes, the stimulating home environment, and maternal mental health. Future work to determine the mechanisms underlying these intervention effects will further inform the design of early interventions to improve child health and development.

**Trial registration:** Follow-up trial: ClinicalTrials.gov, NCT04443855

Original WASH-Benefits Bangladesh (WASH-B): ClinicalTrials.gov, NCT01590095

## Introduction

In low- and middle-income countries (LMICs) it is estimated that one-third of three and four-year-olds fail to meet basic milestones in cognitive or socioemotional development.[1] Early measures of motor, cognitive, and socioemotional development are predictive of later life outcomes, including educational attainment, economic earnings, and socio-emotional behaviour in early adulthood.[2–4] During early childhood, the brain undergoes rapid structural and functional changes; positive experiences are more likely to contribute to the development of synaptic connections important for optimal developmental trajectories and resilience, and negative experiences can shift a child off the optimal developmental trajectory.[5] Thus, interventions to reduce risk factors for impaired development in early life are critical to promoting positive developmental trajectories and later life outcomes.

Similar to classification of nutrition interventions, interventions targeting child development can be classified as development-specific interventions (i.e. those addressing immediate determinants of nutrition and child development, such as inadequate nutrient intake and unsupportive caregiving practices), or development-sensitive interventions (i.e. those addressing the underlying causes of undernutrition or poor development such as poverty, food insecurity, or poor water and sanitation).[6] Development-specific interventions focused on teaching caregivers about the developmental importance of responsive caregiver-child interactions or the provision of nutrition supplements or education have been shown to impact immediate early child development outcomes.[7,8]

Development-sensitive interventions include those that provide nutritional supplementation in early childhood, as well as those that aim to improve water, sanitation, and handwashing (WASH) conditions. Nutrition interventions, which provide the nutrients required for brain development have been shown to improve child development outcomes early in life when delivered in the first 1000 days.[8] WASH interventions aim to reduce the burden of enteric pathogens in the environment. Enteric infections, including intestinal worms caused by poor sanitation, can result in iron deficiency, and have negative consequences for child development outcomes.[9] Observational studies have demonstrated associations between diarrhoea or other infectious diseases and impaired cognitive outcomes.[10] Though observational research demonstrates associations between improved water and sanitation infrastructure and early child development outcomes,[11] few randomized-controlled trials evaluated the independent effects of early WASH interventions on child development outcomes. Three interventions, including the one followed up as part of this study have evaluated the immediate post intervention impact of development-sensitive interventions targeting improvements in WASH on child development outcomes.[12–14] An early WASH intervention in rural Zimbabwe found no effect on child development outcomes. [12] Individual or combined WASH and nutrition interventions improved early child development in Bangladesh, but not in Kenya.[13,14]

Despite robust evidence for the early effects of both development-specific interventions, very few studies have investigated the medium or long-term impacts of early interventions to promote child development on outcomes in middle and late-childhood.[15] There is even less evidence for the medium- or long-term effects of development-sensitive interventions. A follow-up of the early WASH intervention in rural Zimbabwe that had no impact on development outcomes in early childhood found no impacts on cognitive development at 7 years of age, and small impacts of WASH on socioemotional function.[16] Another cRCT in Pakistan found that children whose households received a handwashing promotion and drinking water treatment for 9 months in the first 30 months of life had improved child development at 5-7 years of age.[17] Our team was unable to find any studies that evaluated the later impact of an early combined Nutrition and WASH intervention that previously demonstrated an early impact on child development outcomes.

Our previous cluster-randomized controlled trial (cRCT) in Bangladesh evaluated the impact of water quality, handwashing, sanitation (WASH), and nutrition interventions, when delivered either individually or in combination during pregnancy through 18 months after birth, on diarrhoea, growth, and developmental outcomes in children.[13,18] It was hypothesized that the WASH intervention had the potential to positively benefit the developmental trajectories of children by reducing enteric infection, improving child nutritional status and health, and altering parental interaction and care practices. At the immediate post-intervention evaluation at the age of approximately 2-years, there were immediate benefits on one or more domains of child development in all intervention arms, with the greatest benefit in the combined intervention arm (WASH and nutrition), with effect sizes ranging from 0·13 to 0·35 SDs.[13] Additionally, mothers in all intervention groups reported fewer depressive symptoms compared to mothers in the control households (effect sizes ranged between -0·19 to -0·31 SDs). To better understand how early water, sanitation, and hygiene interventions can impact child development over the life course, we aim to evaluate the effects of individual and combined water, sanitation, hygiene, and nutrition interventions on primary outcomes of child development, school achievement, executive functioning, fine motor development, and socio-emotional development, and secondary outcomes of maternal mental health and stimulation in the home environment at 7 years of age.

## Methods

### Original study design and participants

The parent cluster-randomized controlled trial (WASH-Benefits, or WASH-B, ClinicalTrials.gov Identifier: NCT01590095) was conducted in the rural villages of four districts (Gazipur, Kishoreganj, Mymensingh, and Tangail) of central Bangladesh. The parent trial began in 2011 when there were no major water, sanitation, or focused nutrition programmes in the study area. The details about randomization, study design, intervention, methods, and rationale are described elsewhere.[18,19] In brief, pregnant women were randomized to one of six intervention arms: chlorinated drinking water (W); improved sanitation (S); handwashing with soap (H); combined water, sanitation, and handwashing (WASH); improved nutrition through counselling and provision of lipid-based nutrient supplements (N); and combined water, sanitation, handwashing, and nutrition (WASH+N) or into a double-sized, passive, control arm (C) through geographically pair-matched randomization (Table S1). The intervention continued for two years. A total of 5,551 pregnant women were enrolled, among them, 4,757 live births were assessed at 1 year and 4,403 at 2 years of age.[13]

### Procedure at long term follow-up

Between September 2019 and February 2021, 7 years following intervention initiation, and 5 years following intervention completion, we re-visited the households of all live births who had not been reported to have passed away at any previous time point. Trained enumerators evaluated children, mothers, and the home environment through two separate household visits around a week apart. The visits took 1.5 and 2.5 hours, respectively. During the initial visit (Day-1), trained enumerators obtained written consent and collected information on sociodemographic factors, and data regarding-water supply, sanitation, faecal disposal methods, hand washing conditions, latrine infrastructure, and other facilities. Additionally, they evaluated the home environment, children’s behaviour, and anthropometry. In the subsequent visit (Day-2), enumerators assessed the developmental outcomes of the children and evaluated the depressive symptoms of their mothers.

On Day-1, 10 university graduate enumerators conducted data collection and assessments. On Day-2, 12 enumerators, who had degrees in social sciences or psychology and experiences in child development assessments, conducted developmental evaluations. The Day-1 and Day-2 enumerators underwent a 2 (for day 1) and 4 (for day 2) training programs, which included theoretical instruction, mock practice, and hands-on practice with non-study participants/children in the community, under supervision. For Day-2 enumerators inter-observer reliability assessments were conducted between testers and enumerators where each enumerator conducted at least 10 tests. Enumerators were only allowed to participate in the main study once they achieved over 90% agreement with the trainers. During the data collection phase, a supervisor evaluated 10% of all tests to ensure satisfactory ongoing reliability (kappa > 0.90). Refresher training sessions were held once in October 2020 for both data collection teams. Enumerators were blinded to intervention status. Up to three attempts were made to find each household. Children who were vision or hearing impaired or had a severe developmental disability were excluded from the assessment.

The study protocol was approved by human subjects committees at icddr,b (PR-19025), and the University of California, Berkeley (2018-12-11672).

### Measurement

#### Primary outcomes

To measure child cognitive development, we selected 9 subtests of the Wechsler Pre and Primary School Intelligence – Fourth Edition (WPPSI-IV) developed in the United States.[20] This test aims to assess the intellectual ability of children aged 4 to 7.5 years of age. Previous versions of WPPSI have been widely implemented in Bangladesh, and went through rigorous cultural adaptations before use.[21] Our cultural adaptations focused on modifying individual items and pictures that were unfamiliar in the rural Bangladeshi context (e.g. we replaced the image of “Red Fire Hydrant” with “local Red Gas Cylinder”, “Life Jacket” with “Normal Jacket”, “Hour Glass” with “Table-Clock”) while maintaining the underlying intent of the question. We followed standard instructions for administration and scoring of evaluations. Using the 9 subtests we constructed subscales for the Full Scale IQ (FSIQ); 3 Primary Index Scores: Verbal Comprehension Index (VCI), Fluid Reasoning Index (FRI), Working Memory Index (WMI); 3 Ancillary Index Score: General Ability Index (GAI), Nonverbal Index (NVI), and the Cognitive Proficiency Index (CPI).

To assess children’s fine motor ability we used three measures of manual dexterity following the second edition of the Movement Assessment Battery for Children.[22] These tests have been previously used in Bangladesh.[23] To assess children’s behaviour we used the parent-reported Strengths and Difficulties Questionnaire (SDQ).[24] The standard Bengali language (Bangla) version of SDQ has been used previously in Bangladesh.[25] We assessed three areas of children’s executive functioning. Working memory, attention, and memory (short and delayed) were evaluated using forward Digit span, Corsi-blocks, and a narrative memory test, respectively. These tests were chosen from neuropsychological[26] and preschool assessment tools[27] after piloting on 50 children. We piloted a battery of executive function tests previously implemented in South Asia and selected the tests that showed variability for children of the age group assessed in the current study. Finally, we evaluated academic achievement (reading, spelling, and mathematics) using a locally developed tool based on the Wide Range Achievement Test (WRAT).[28] It has individually administered reading, spelling, and arithmetic questions in Bengali which are ranked with increasing difficulty. It has previously been used in Bangladesh with primary school children.[29]

We internally age-standardized all primary outcomes to the control group separately for data collected prior to the COVID pandemic and data collected during the COVID pandemic. We used local mean standardization with 2-month age bands. We had originally planned to use 4-month age bands but had sufficient sample size to reduce the size of the age band. To construct WPPSI-IV and fine motor subtest scores, we first internally standardized each subscale, then averaged the internally standardized scores across relevant subtest for each subscale, and finally constructed a z-score with respect to the control group. We conducted supplementary analyses with externally standardized scores for the WPPSI-IV subscales.

#### Secondary outcomes

We measured maternal depressive symptoms of mothers with the 20-item Center for Epidemiologic Studies Depression Scale (CESD).[30] The 20-item CESD has been used widely in Bangladesh and other South-East Asian countries.[31] We evaluated the home environment and amount of stimulation at home with an adapted version of the Middle childhood Home Observation Measurement (HOME), which has been used previously in Bangladesh.[29,32]

### Other measurements

We collected information on the sustained presence of technologies distributed during the initial intervention following protocols used at the 1 and 2 year follow up time points.[18]

### Statistical Analysis

The sample size for the original trial was calculated to detect a difference of 0.15 in the length-for-age Z score (LAZ) when comparing each intervention arm to the control, accounting for repeated measures within clusters.[19] In this follow-up, for each outcome, we compared the control arm to each intervention arm, and also compared the WSH+N vs. Nutrition, and WSH+N vs WSH to isolate the additive effects of WSH and N, respectively. We first calculated unadjusted mean differences using generalized linear models accounting for pair matching and block-level clustering, to account for the original randomization procedure.[18] We then calculated adjusted mean differences controlling for time of assessment (pre-COVID or during COVID) and concurrent child age along with a set of prognostic baseline characteristics. The covariates for potential inclusion in each model were assessed with a likelihood ratio test, and covariates with p>0.20 were included. All analyses were intention to treat.

We conducted subgroup analyses by maternal primary education status at baseline, child sex at birth, measurement before or during the COVID-19 pandemic, and socioeconomic status at baseline (wealth index split at the median). and We had originally planned to conduct additional stratified analyses by household distance from Dhaka (split at the median distance) but found that there was large overlap between distance from Dhaka and measurement pre vs. post COVID, so we did not conduct this analysis.

All analyses were done in R, version 4.3.3.[33] The pre-analysis plan for this study is posted on OSF (https://osf.io/jgh7y/?view_only=ca62a737fc3a443f859e83eb6cc9ad13). In the pre-analysis plan, we specified that we would additionally do subgroup analysis by child age at assessment, but the lack of full age overlap across assessment periods made this not viable as an independent analysis.

The follow-up is registered with ClinicalTrials.gov, NCT04443855.

## Results

Between September 2019 and February 2021, enumerators attempted to locate 4,961 children in 4,932 households (Figure 1). Assessments were paused during the initial phases of the COVID-19 pandemic from March 15^th^ to October 13^th^, 2020. The assessment took place over two days, which aimed to be conducted within 3 weeks.

**Figure 1.**
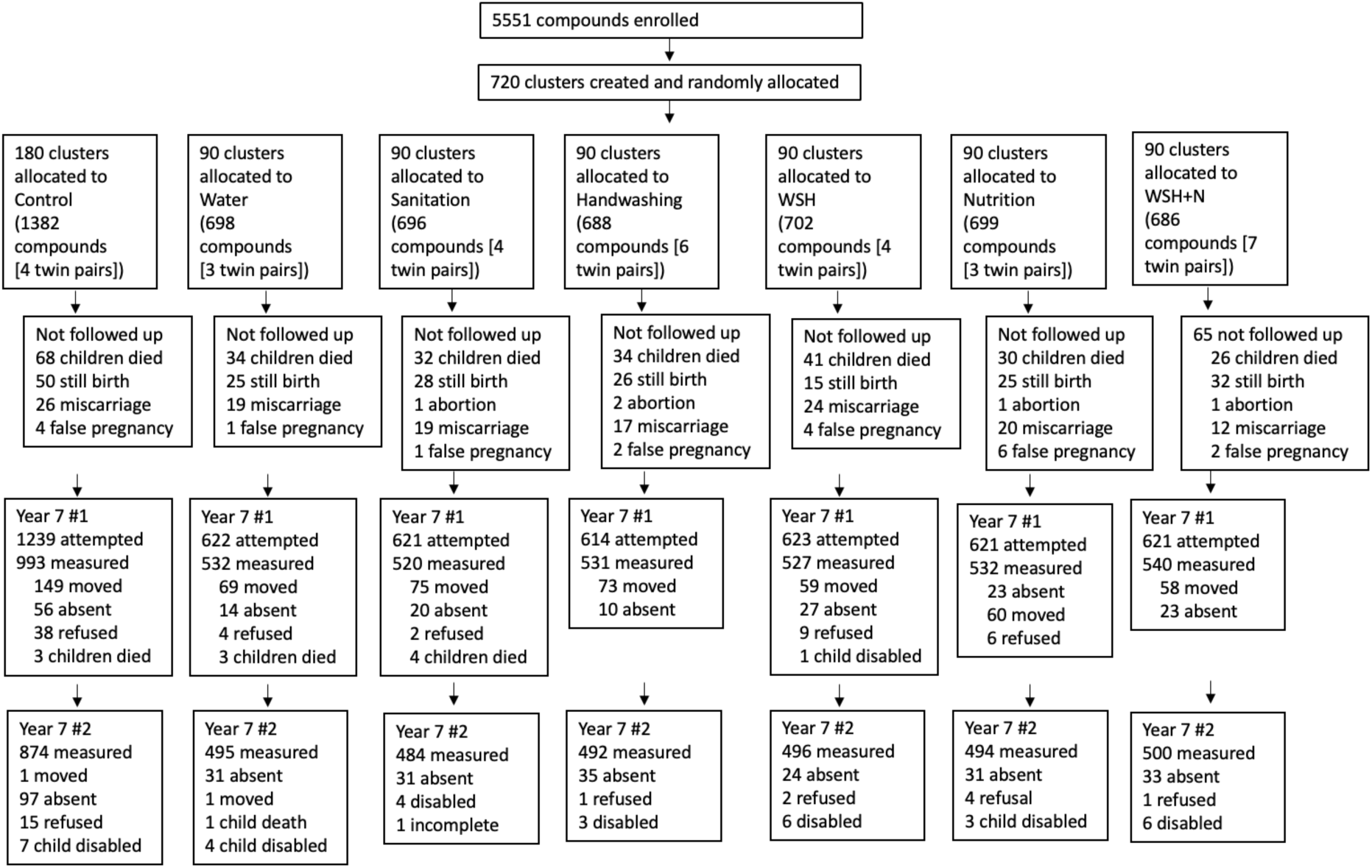
Flow diagram. Follow up #1: Included water, sanitation and hygiene infrastructure, school attendance information, HOME assessment, Strengths and difficulties questionnaire, anthropometry; Follow up #2: included all direct cognitive, motor and school achievement assessments, and maternal mental health.

Due to COVID-19 pandemic related disruptions as well as households that were unavailable for a second assessment within 3 weeks, 23% of assessments were conducted over 3 weeks apart. A total of 4,175 (84% of those attempted) children completed the first assessment, with the most common reasons for not completing the assessment were that the family had migrated (n=543, 11% of those attempted) or the child was absent from household (n=173, 3%) (Figure 1). At the second assessment, which included the child development assessment, an additional 340 (8% of the sample attempted) did not complete the assessment, with the most common reason being that the child was absent from the household at the second visit (n=281, 7% of those attempted for visit 2) (Figure 1). Complete assessments for both days were performed for 3,833 (69% of the initially enrolled sample, 77% of the sample that was attempted at follow-up) children in 3,812 households (21 pairs of twins) (Figure 1). Participants followed-up were similar to those who were lost to follow-up across a range of characteristics (Table S2).

Baseline characteristics of the sample are similar across study arm for those in the completely assessed group at the 7-year follow-up (Table 1). Children had a mean age of 83.5 months (Range: 74.4-92.2, SD: 3.5) at the child development assessment.

**Table 1.** Baseline characteristics of the re-enrolled populations.

| No. of clusters | Control<br>179<br>(N = 874) | Water<br>90<br>(N = 495) | Sanitation<br>90<br>(N = 484) | Handwashing<br>90<br>(N = 492) | WSH<br>90<br>(N = 496) | Nutrition<br>89<br>(N = 494) | WSH+N<br>90<br>(N = 500) |
| --- | --- | --- | --- | --- | --- | --- | --- |
| **Maternal** |  |  |  |  |  |  |  |
| Age (years) | 24.0 (5.0) | 24.0 (5.3) | 24.2 (5.0) | 23.9 (5.4) | 24.7 (5.7) | 23.9 (5.2) | 24.1 (5.5) |
| Years of education | 5.7 (3.4) | 5.7 (3.4) | 6.0 (3.5) | 5.9 (3.3) | 5.9 (3.3) | 5.8 (3.5) | 5.6 (3.5) |
| **Paternal** |  |  |  |  |  |  |  |
| Years of education | 4.8 (3.9) | 4.8 (4.0) | 5.2 (4.1) | 4.7 (4.1) | 5.0 (4.2) | 4.7 (3.9) | 4.7 (3.9) |
| Works in agriculture | 283 (32%) | 166 (34%) | 150 (31%) | 190 (39%) | 161 (32%) | 164 (33%) | 148 (30%) |
| **Household** |  |  |  |  |  |  |  |
| Number of persons | 4.7 (2.1) | 4.7 (2.3) | 4.8 (2.1) | 4.8 (2.2) | 4.6 (2.0) | 4.6 (2.0) | 4.8 (2.1) |
| Has electricity | 519 (59%) | 305 (62%) | 297 (61%) | 292 (59%) | 319 (64%) | 299 (61%) | 301 (60%) |
| Has a cement floor | 96 (11%) | 56 (11%) | 61 (13%) | 37 (8%) | 56 (11%) | 45 (9%) | 61 (12%) |
| Acres of agricultural land owned | 0.1 (0.2) | 0.1 (0.2) | 0.2 (0.3) | 0.2 (0.2) | 0.2 (0.2) | 0.2 (0.3) | 0.1 (0.4) |
| **Drinking Water** |  |  |  |  |  |  |  |
| Tubewell primary water source | 670 (77%) | 360 (73%) | 362 (75%) | 341 (69%) | 389 (78%) | 370 (75%) | 364 (73%) |
| Stored water observed at home | 422 (48%) | 249 (50%) | 230 (48%) | 242 (49%) | 217 (44%) | 212 (43%) | 236 (47%) |
| **Sanitation** |  |  |  |  |  |  |  |
| Daily defecation in the open |  |  |  |  |  |  |  |
| Adult men | 63 (7%) | 24 (5%) | 33 (7%) | 46 (9%) | 31 (6%) | 37 (8%) | 38 (8%) |
| Adult women | 46 (5%) | 13 (3%) | 22 (5%) | 21 (4%) | 18 (4%) | 23 (5%) | 18 (4%) |
| Children: 3-<8 years | 190 (40%) | 104 (37%) | 103 (39%) | 99 (38%) | 93 (36%) | 92 (38%) | 94 (35%) |
| Children: 0-<3 years | 157 (84%) | 82 (87%) | 81 (83%) | 94 (84%) | 82 (78%) | 82 (80%) | 87 (91%) |
| **Latrine** |  |  |  |  |  |  |  |
| Owned | 452 (52%) | 263 (53%) | 262 (54%) | 271 (55%) | 263 (53%) | 266 (54%) | 269 (54%) |
| Concrete slab | 785 (94%) | 461 (96%) | 427 (92%) | 443 (94%) | 451 (94%) | 445 (94%) | 454 (94%) |
| Functional water seal | 226 (31%) | 136 (31%) | 126 (31%) | 124 (30%) | 104 (25%) | 135 (32%) | 113 (27%) |
| Visible stool on slab or floor | 406 (49%) | 252 (53%) | 236 (53%) | 239 (52%) | 209 (45%) | 229 (50%) | 220 (47%) |
| Owned a potty | 32 (4%) | 17 (3%) | 19 (4%) | 23 (5%) | 20 (4%) | 28 (6%) | 18 (4%) |
| Human feces observed in the |  |  |  |  |  |  |  |
| House | 79 (9%) | 47 (10%) | 43 (9%) | 54 (11%) | 32 (6%) | 40 (8%) | 33 (7%) |
| Child's play area | 12 (1%) | 6 (1%) | 6 (1%) | 7 (1%) | 3 (1%) | 5 (1%) | 5 (1%) |
| **Handwashing** |  |  |  |  |  |  |  |
| Within 6 steps of latrine |  |  |  |  |  |  |  |
| Has water | 115 (14%) | 63 (14%) | 59 (13%) | 42 (10%) | 42 (9%) | 44 (10%) | 60 (13%) |
| Has soap | 58 (7%) | 34 (8%) | 34 (8%) | 23 (5%) | 25 (6%) | 24 (5%) | 27 (6%) |
| Within 6 steps of kitchen |  |  |  |  |  |  |  |
| Has water | 75 (9%) | 32 (7%) | 35 (8%) | 25 (6%) | 40 (9%) | 45 (10%) | 45 (10%) |
| Has soap | 21 (3%) | 10 (2%) | 11 (2%) | 6 (1%) | 11 (2%) | 17 (4%) | 15 (3%) |
| **Nutrition** |  |  |  |  |  |  |  |
| Food secure | 595 (68%) | 352 (71%) | 335 (69%) | 342 (70%) | 337 (68%) | 337 (68%) | 356 (71%) |

Data are n (%) or mean (s.d.); No. of participants includes 21 sets of twins

At the 7-year follow-up, households in intervention arms that received the sanitation intervention were more likely to have a latrine with a functional water seal (Sanitation: 83%, WSH: 78%, and WSH+N: 83%) compared to the control arm (48%) and other arms without sanitation (Sanitation: 51%, WSH: 50%, WSH+N: 51%) (Table 2). The proportion of households in the Sanitation arms with a latrine with a functional water seal was lower than at the 2-year endline (Table 2). Other indicators of intervention adherence including the presence of stored drinking water, visible faeces on the latrine slab or floor, and the presence of soap at the handwashing station were not substantially different across arms (Table 2).

**Table 2.**
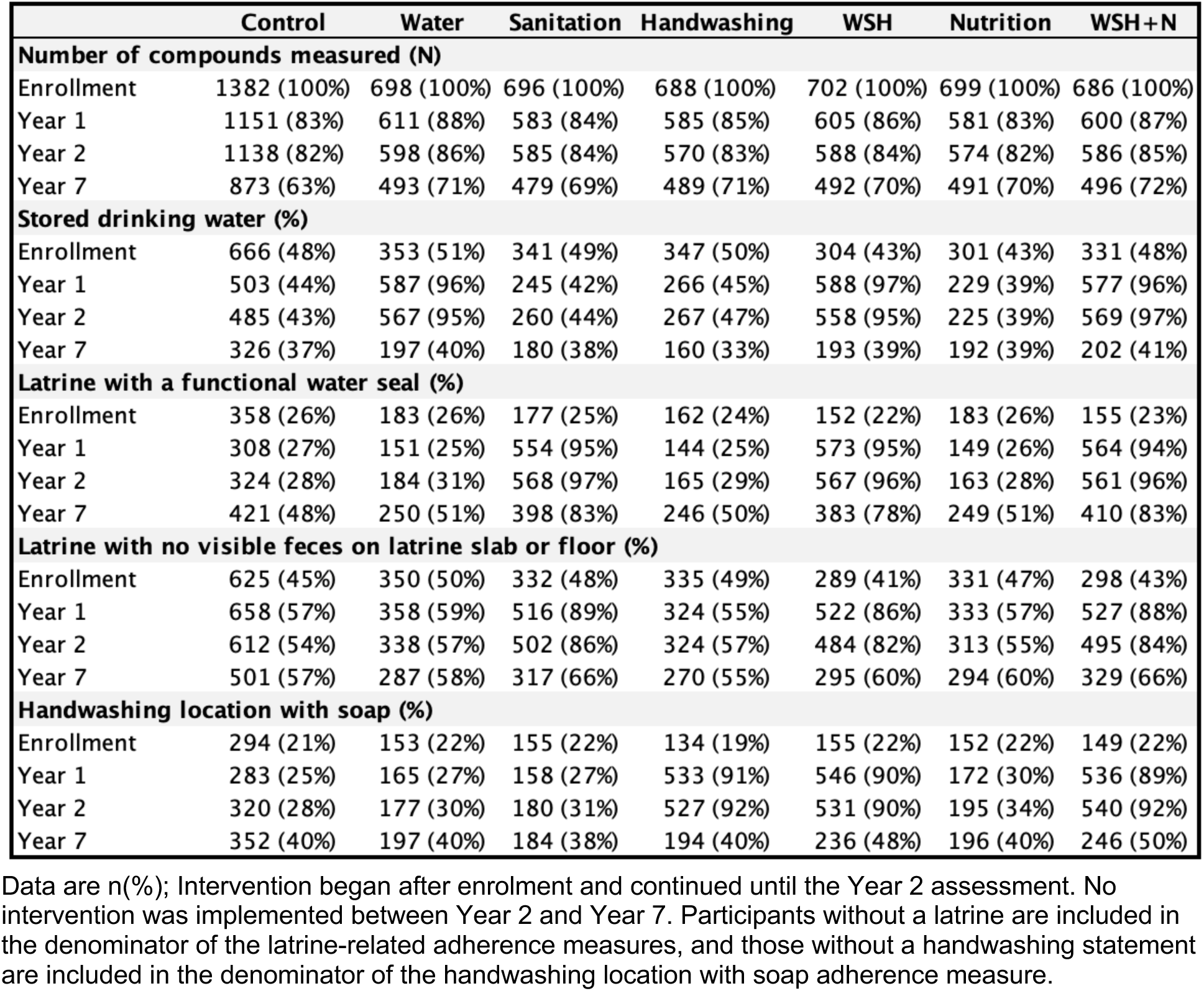
Sustained adoption of behavioural recommendations and continued presence of technologies promoted or distributed during the period of project implementation.

|  | Control | Water | Sanitation | Handwashing | WSH | Nutrition | WSH+N |
| --- | --- | --- | --- | --- | --- | --- | --- |
| **Number of compounds measured (N)** |  |  |  |  |  |  |  |
| Enrollment | 1382 (100%) | 698 (100%) | 696 (100%) | 688 (100%) | 702 (100%) | 699 (100%) | 686 (100%) |
| Year 1 | 1151 (83%) | 611 (88%) | 583 (84%) | 585 (85%) | 605 (86%) | 581 (83%) | 600 (87%) |
| Year 2 | 1138 (82%) | 598 (86%) | 585 (84%) | 570 (83%) | 588 (84%) | 574 (82%) | 586 (85%) |
| Year 7 | 873 (63%) | 493 (71%) | 479 (69%) | 489 (71%) | 492 (70%) | 491 (70%) | 496 (72%) |
| **Stored drinking water (%)** |  |  |  |  |  |  |  |
| Enrollment | 666 (48%) | 353 (51%) | 341 (49%) | 347 (50%) | 304 (43%) | 301 (43%) | 331 (48%) |
| Year 1 | 503 (44%) | 587 (96%) | 245 (42%) | 266 (45%) | 588 (97%) | 229 (39%) | 577 (96%) |
| Year 2 | 485 (43%) | 567 (95%) | 260 (44%) | 267 (47%) | 558 (95%) | 225 (39%) | 569 (97%) |
| Year 7 | 326 (37%) | 197 (40%) | 180 (38%) | 160 (33%) | 193 (39%) | 192 (39%) | 202 (41%) |
| **Latrine with a functional water seal (%)** |  |  |  |  |  |  |  |
| Enrollment | 358 (26%) | 183 (26%) | 177 (25%) | 162 (24%) | 152 (22%) | 183 (26%) | 155 (23%) |
| Year 1 | 308 (27%) | 151 (25%) | 554 (95%) | 144 (25%) | 573 (95%) | 149 (26%) | 564 (94%) |
| Year 2 | 324 (28%) | 184 (31%) | 568 (97%) | 165 (29%) | 567 (96%) | 163 (28%) | 561 (96%) |
| Year 7 | 421 (48%) | 250 (51%) | 398 (83%) | 246 (50%) | 383 (78%) | 249 (51%) | 410 (83%) |
| **Latrine with no visible feces on latrine slab or floor (%)** |  |  |  |  |  |  |  |
| Enrollment | 625 (45%) | 350 (50%) | 332 (48%) | 335 (49%) | 289 (41%) | 331 (47%) | 298 (43%) |
| Year 1 | 658 (57%) | 358 (59%) | 516 (89%) | 324 (55%) | 522 (86%) | 333 (57%) | 527 (88%) |
| Year 2 | 612 (54%) | 338 (57%) | 502 (86%) | 324 (57%) | 484 (82%) | 313 (55%) | 495 (84%) |
| Year 7 | 501 (57%) | 287 (58%) | 317 (66%) | 270 (55%) | 295 (60%) | 294 (60%) | 329 (66%) |
| **Handwashing location with soap (%)** |  |  |  |  |  |  |  |
| Enrollment | 294 (21%) | 153 (22%) | 155 (22%) | 134 (19%) | 155 (22%) | 152 (22%) | 149 (22%) |
| Year 1 | 283 (25%) | 165 (27%) | 158 (27%) | 533 (91%) | 546 (90%) | 172 (30%) | 536 (89%) |
| Year 2 | 320 (28%) | 177 (30%) | 180 (31%) | 527 (92%) | 531 (90%) | 195 (34%) | 540 (92%) |
| Year 7 | 352 (40%) | 197 (40%) | 184 (38%) | 194 (40%) | 236 (48%) | 196 (40%) | 246 (50%) |

Data are n(%); Intervention began after enrolment and continued until the Year 2 assessment. No intervention was implemented between Year 2 and Year 7. Participants without a latrine are included in the denominator of the latrine-related adherence measures, and those without a handwashing statement are included in the denominator of the handwashing location with soap adherence measure.

### WPPSI-IV

Compared to children in the control arm, children in the WSH+N arm had better FSIQ scores (adjusted mean difference: 0.12 (95% CI: 0.02, 0.23)) (Figure 2, Table S3). Children in all other arms did not have differences in FSIQ scores compared to children in the control arm that were statistically significant at p<0.05 (Figure 2, Table S3). Compared to children in the control arm, children had better FRI scores in the WSH+N (0.15 (0.03, 0.27)), Sanitation (0.12 (0.00, 0.24)), and Nutrition arms (0.12 (0.01, 0.23)) (Figure S1, Table S3). Children also had better WPPSI-IV GAI scores in the WSH+N (0.12 (0.03, 0.22)), Sanitation (0.12 (0.01, 0.23)), and Nutrition (0.11 (0.01, 0.20)) arms, better NVI scores in the WSH+N (0.13 (0.03, 0.24)) and Sanitation (0.11 (0.00, 0.22)) arm, and better VCI scores in the nutrition arm (0.10 (0.00, 0.20)) (Figure S1, Table S3). There were no differences between any intervention arm and the control arm for the CPI or WMI scores (Figure S1, Table S3).

**Figure 2.**
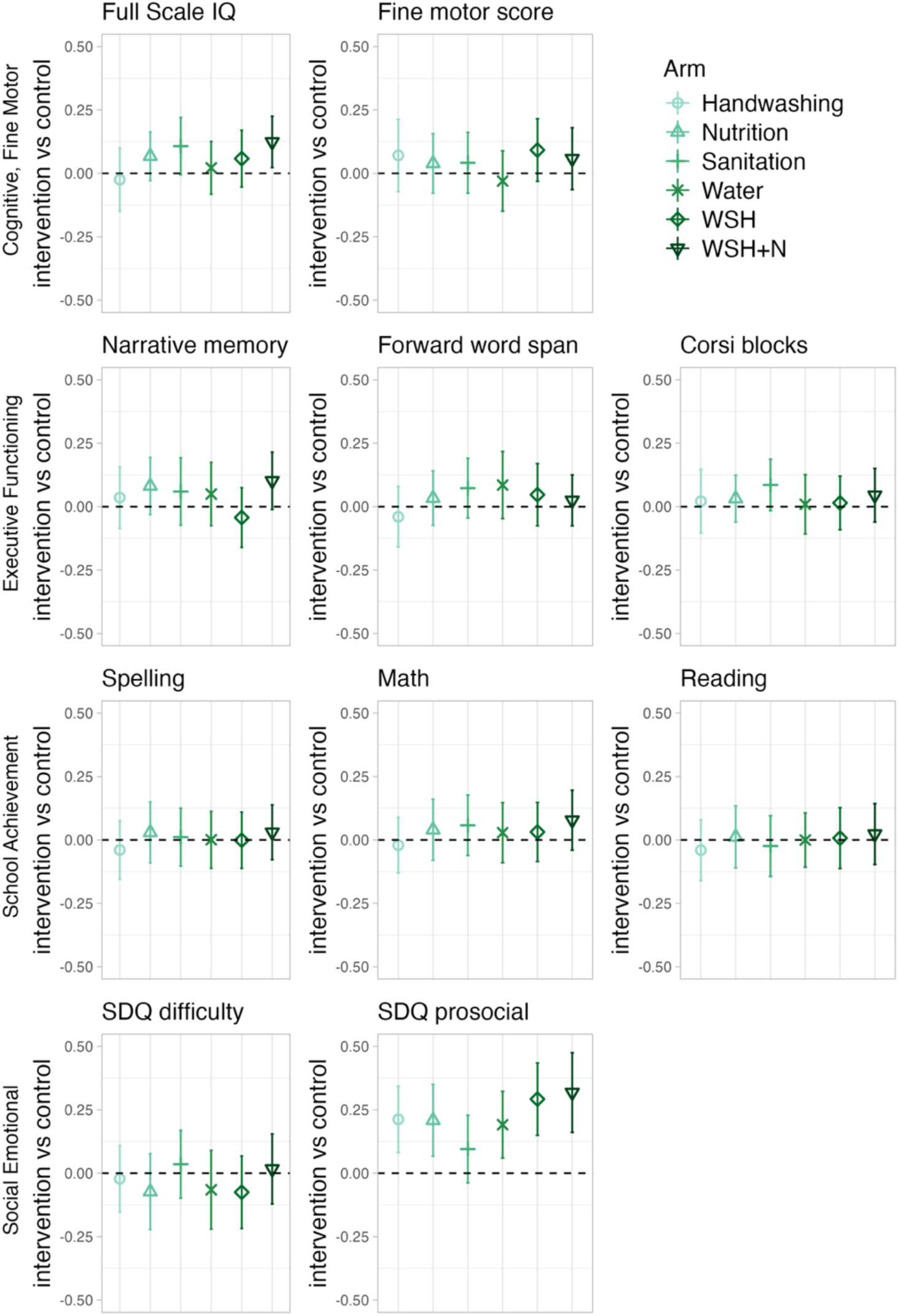
Differences in child development and school achievement by control versus intervention arm. SDQ: strengths and difficulties questionnaire, Narrative memory: Sum of free and cued recall scores from a narrative memory test

### Fine motor, executive functioning, academic achievement, difficult and prosocial behaviour

There were no differences between the control arm and any intervention arm for fine motor development, executive functioning, or the spelling, math, or reading domains of the school achievement test (Figure 2, Table S4, Figure S2). The measure of social emotional development showed that difficult behaviours were not different between any intervention arm when compared to the control arm, but prosocial behaviour scores were higher for children in all intervention arms except for the sanitation arm compared to the control arm (Water: 0.19 (0.06, 0.32), Hygiene: 0.21 (0.08, 0.34), WSH: 0.29 (0.15, 0.43), Nutrition: 0.21 (0.07, 0.35), WSH+N: 0.32 (0.16, 0.48) (Figure 2, Table S4).

Compared to caregivers of control children, caregivers had fewer depressive symptoms in the WSH+N (-0.15 (-0.27, -0.02)), Handwashing (-0.14 (-0.24, -0.03)), and Nutrition (-0.21 (-0.31, -0.11)) arms. Compared to children in the control arm, children in all intervention arms had more stimulating home environments ((Water: 0.19 (0.03, 0.35), Sanitation: 0.17 (0.00, 0.35), Hygiene: 0.26 (0.11, 0.42), WSH: 0.31 (0.15, 0.48), Nutrition: 0.24 (0.08, 0.41), WSH+N: 0.41 (0.24, 0.57)) (Figure 3, Table S5).

**Figure 3.**
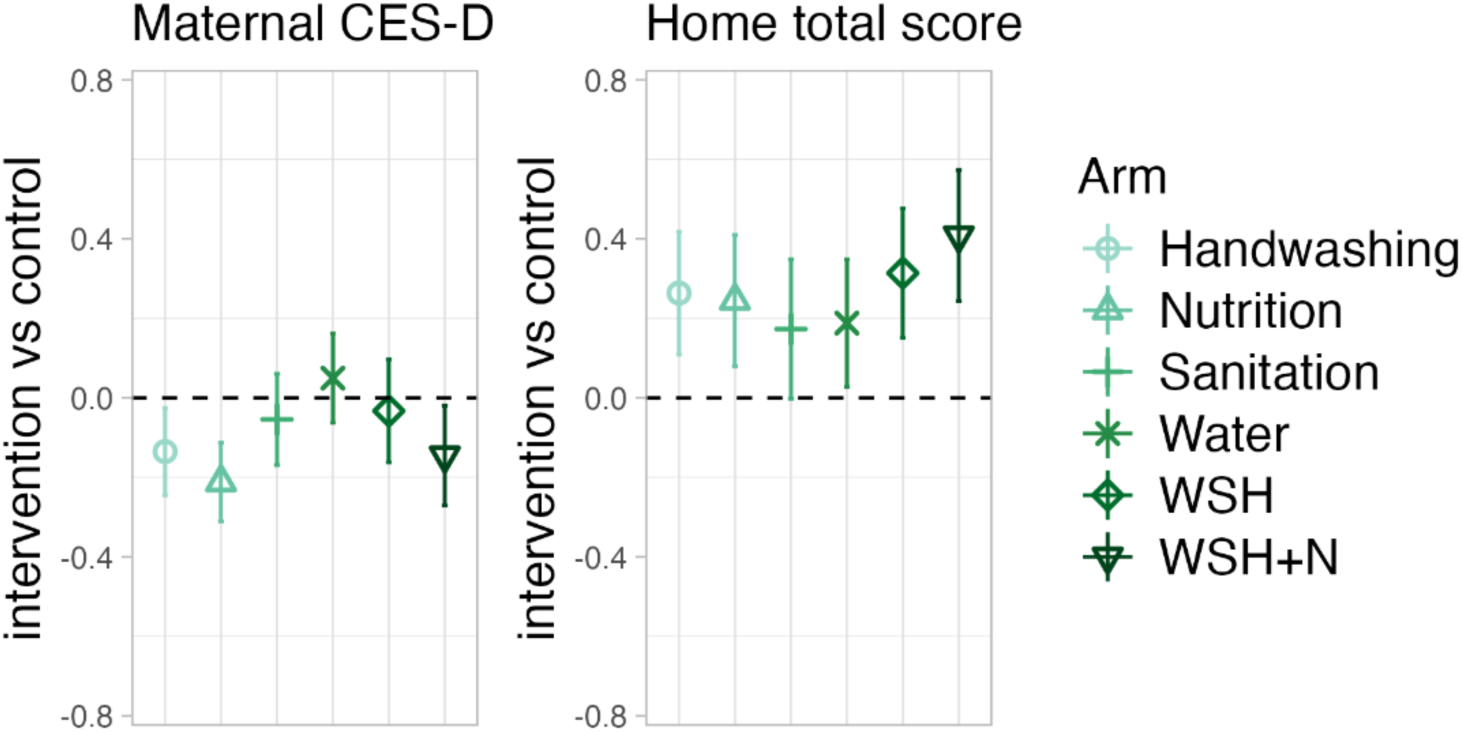
Differences in secondary outcomes by arm. CES-D: Center for epidemiologic studies 20-question depression measure; HOME: Middle childhood Home Observation Measurement of the Environment

In subgroup analyses there was no consistent heterogeneity in intervention effects on FSIQ by assessment timing (prior to or during the COVID pandemic), or maternal education status (mother had completed secondary school prior to pregnancy) (Figure 4). Families with higher wealth indices at baseline, and those with male children tended to have larger effect sizes on the FSIQ (Figure 4). There were no consistent patterns across other outcomes for heterogeneity analyses (Figures S3-S12). There were no differences between WSH+N vs nutrition or WSH+N vs WSH for any outcomes (Tables S1-S4).

**Figure 4.**
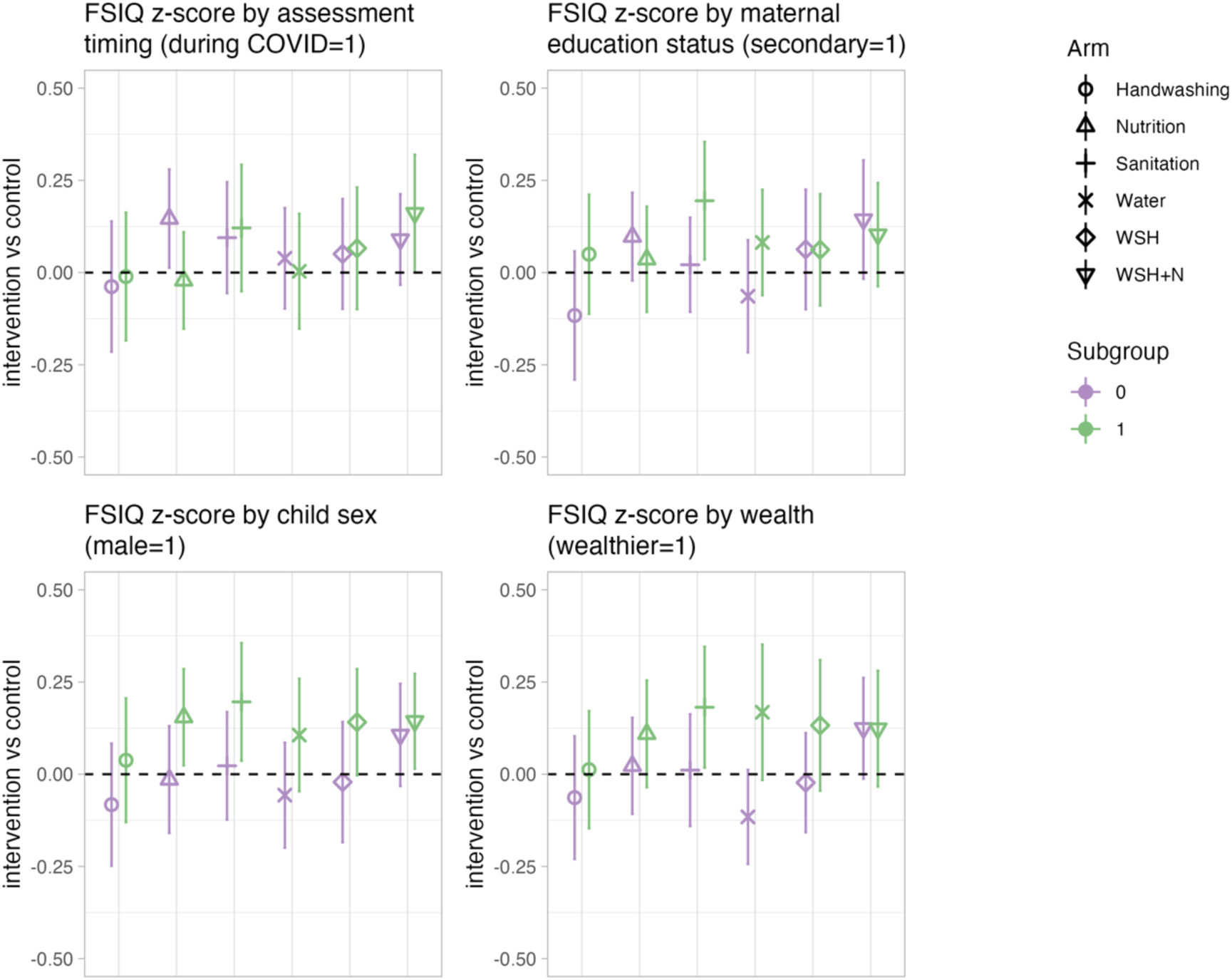
Subgroup analysis for FSIQ outcome. FSIQ: Full Scale IQ

## Discussion

This medium-term follow up showed that 5 years after intervention completion single or combined water, handwashing, sanitation, and nutritional interventions had small but significant sustained impacts on one or more domains of child development, the home environment, and caregiver mental wellbeing. At approximately 7 years of age, we found that children who received the WSN+N, Nutrition, or Sanitation intervention had improved cognitive development in three of the six WPPSI-IV subscales when compared to control children, and children in all arms except for the Sanitation arm had improved prosocial behaviours. Children in all intervention arms had more stimulating home environments compared to the control arm. Caregiver-related outcomes were also improved, with caregivers in the WSH+N, Handwashing, and Nutrition arms having fewer depressive symptoms. There were no improvements in fine motor, executive functioning or academic achievement for children in any of the intervention arms. Impacts across domains did not substantially differ by assessment timing (prior to or during the COVID-19 pandemic), maternal education status, child sex, or wealth status.

Our intervention was unique in that it followed up an early WASH intervention that improved child development at 2 years of age. Two other randomized controlled trials have evaluated the impact of early WASH interventions on early child development outcomes immediately post intervention, and neither found any improvements for children who received the interventions. [12,14] One previous study in Pakistan evaluated later, but not immediate impacts of a handwashing and water intervention on child development. In Pakistan, children who were randomized to receive either hand washing promotion or hand washing promotion plus water treatment interventions during the first 30 months of life were followed up between 5-7 years of age. Children who received the intervention were found to have improvements on a composite measure of development equivalent to an effect size of 0.4 standard deviations.[17] Other interventions have evaluated a combination of sanitation or hygiene interventions with stimulation on outcomes later in childhood, and found improved child development and caregiver mental health amongst families who received the intervention.[34,35] However, these interventions are unable to untangle the impact of the water, sanitation and hygiene component from the psychosocial stimulation component.

In contrast to the evidence for early WASH interventions, there is evidence for the impact of early nutrition supplementation on immediate child development. A recent systematic review and meta-analyses,[8] examined the impact of early small-quantity lipid-based multiple micronutrient supplements delivered within the first 1000 days on short-term child-development outcomes in LMICs. They found that 11 out of 13 interventions showed moderate post-intervention benefits to language, social-emotional, motor, and executive function. However, few of these studies followed up with children. In rural Pakistan a nutrition education and multiple micronutrient powder supplementation intervention for children 6-24 months of age was found to improve motor development, but not WPPSII-III FSIQ, executive functioning, pre-academic skills or behavioural problems at 4 years of age.[36] In Ghana, lipid-based nutrient supplements provided between 6-24 months reduced social emotional difficulties amongst children at 4-6 years of age, but weren’t found to impact cognitive or fine motor development outcomes.[37] These impacts on social emotional difficulties did not persist to 9-11 years of age.[38] A micronutrient supplementation trial in Indonesia found improved procedural memory in children at 9-12 years of age, and enrolled children born to anaemic mothers had improved general intellectual ability.[39] Finally, a small factorial designed nutrition and stimulation intervention provided in Jamaica found no impact of 2 years of early supplementation with milk-based formula on child development outcomes at 7-8 years of age except a subset of children with mothers who had higher verbal intelligence quotients, and no impact was found from supplementation at later time points.[40]

In this follow-up, we found small but sustained benefits in Full Scale IQ for children in the Sanitation and WSH+N arms. We also found improvements in three out of six total WPPSI sub-domains for the WSH+N, Nutrition, and Sanitation arms, and improved prosocial behaviours in all intervention arms. The intervention was given during the first 1000 days of life, a critical and sensitive window of development.[5,41] There are two primary pathways through which we hypothesize these interventions could have had sustained impacts. The first is through reduced enteric infections, as the initial interventions reduced diarrhoea at 2 years of age in all intervention arms except for the Water arm.[18] Inflammation associated with diarrhoea limits the absorption of key nutrients, and repeated episodes of diarrhoea before age two are associated with poorer cognitive development outcomes, so this may also be a mechanism by which the early intervention had sustained effects.[42,43] Additionally, the two previous WASH interventions that did not find impacts on child development at intervention completion also did not find impacts on diarrhoea.[44,45] Another potential pathway of intervention effect could be through support of caregivers and improved caregiving practices, which was higher in the WASH-Benefits Bangladesh intervention than in the previous WASH interventions. In all intervention arms community health promotors were instructed to visited intervention households weekly for the first 6 months, and then every 2 weeks for the subsequent 18 months, the actual number of visits per month were an average of 5-7 throughout the intervention period.[46] These visits started during the 1^st^ and 2^nd^ trimester of pregnancy and continued for almost two years after birth. Although support for caregiver mental wellbeing or child stimulation was not explicitly provided, maternal depressive symptoms were significantly reduced in the WSH+N, H, and N arms, and the stimulating home environment was improved in all intervention arms. Caregiver mental health and the caregiving environment were also improved at the 2-year endline assessment reported previously.[13] The impacts on child development may be due to increased attention to the child and social support provided to the caregiver during the frequent household visits leading to both improved mental health of caregivers and more attention to and investment in the child’s physical and social environment. Improvements in home stimulation is a target of many child development-specific interventions.[47]

While we found impacts on some measures of child development, for multiple measures we did not identify any impacts. For example, we found impacts on narrative memory for children in the WSH+N arm, but did not find any impacts for any other intervention arms or any other measures of executive functioning. To evaluate executive functioning, we chose tests that focused on memory and attention but did not include a test that evaluated inhibitory control as it was not adequately adapted to the population (we observed ceiling effects during piloting). The chosen tests may not have been sensitive enough to evaluate the intervention’s impact. We also did not find impacts on fine motor, problematic behaviours, or academic achievement. A lack of effects on school achievement may be because most children have only been exposed to one fewer years of formal schooling at this age. As intervention effects did not differ between children assessed prior to and during the COVID-19 pandemic, educational interruption during the early COVID-19 pandemic period was not likely to have affected this finding.

Our study has several strengths including its basis on a large RCT design with well-balanced intervention arms and a double-sized control. We delivered both individual and combined interventions to understand the unique and combined impacts of each intervention. The enumerators went through a rigorous training for developmental assessments and monitored for 10% quality-check throughout the study period. There were direct assessments for children’s development and observation for home environment, and all analyses were prespecified. Our outcome measures had good psychometric properties. However, this study also has several weaknesses. We applied child development assessment tools that were not initially developed or standardized for use in LMIC settings. We culturally adapted all measures for use in our study setting in Bangladesh and conducted rigorous piloting. To further mitigate this issue, instead of using standardized scores from external high-income populations, we used internally standardized z-scores. Further, our data collection period was interrupted by the COVID-19 pandemic, and paused between March and September 2020, which may have affected the results. We adjusted for assessment time period (before or during the COVID-19 pandemic) within the standardization of outcomes, controlled for it in all analyses, and conducted sub-group analyses to examine differences. Finally, we were unable to assess 31% of children from the original study. However, we did not find major differences in baseline characteristics between participants who lost to follow and those followed up, reducing the potential bias induced by loss to follow up.

We found that an early water, sanitation, hygiene, and nutrition interventions had sustained impacts on child development, caregiver mental health, and the stimulating home environment. Future work to elucidate the mechanisms of these impacts will help to isolate the key components of these interventions.

## Data Availability

All data produced in the present study are available upon reasonable request to the authors

## Abbreviations

CESD: Center for Epidemiological Studies Depression Scale
CPI: Cognitive Processing Index
cRCT: cluster-Randomized Controlled Trial
FSIQ: Wechsler preschool and primary scales of intelligence Full Scale IQ
FRI: Fluid Reasoning Index
GAI: General Abilities Index
LMIC: low- and middle-income country
NVI: Non-verbal index
VCI: Verbal Comprehension Index
WMI: Working Memory index
WASH: Water, Sanitation, and Hygiene

## References

1. McCoy DC, Peet ED, Ezzati M, Danaei G, Black MM, Sudfeld CR, et al. Early Childhood Developmental Status in Low- and Middle-Income Countries: National, Regional, and Global Prevalence Estimates Using Predictive Modeling. PLoS Med. 2016;13: e1002034. doi:10.1371/journal.pmed.1002034

2. Gertler P, Heckman J, Pinto R, Zanolini A, Vermeersch C, Walker S, et al. Labor market returns to an early childhood stimulation intervention in Jamaica. Science. 2014;344: 998–1001. doi:10.1126/science.1251178

3. Peet ED, McCoy DC, Danaei G, Ezzati M, Fawzi W, Jarvelin M-R, et al. Early Childhood Development and Schooling Attainment: Longitudinal Evidence from British, Finnish and Philippine Birth Cohorts. PLOS ONE. 2015;10: e0137219. doi:10.1371/journal.pone.0137219

4. Black MM, Walker SP, Fernald LCH, Andersen CT, DiGirolamo AM, Lu C, et al. Early childhood development coming of age: science through the life course. The Lancet. 2017;389: 77–90. doi:10.1016/S0140-6736(16)31389-7

5. Bhutta ZA, Guerrant RL, Nelson CA. Neurodevelopment, Nutrition, and Inflammation: The Evolving Global Child Health Landscape. Pediatrics. 2017;139: S12–S22. doi:10.1542/peds.2016-2828D

6. Ruel MT, Alderman H. Nutrition-sensitive interventions and programmes: how can they help to accelerate progress in improving maternal and child nutrition? The Lancet. 2013;382: 536–551. doi:10.1016/S0140-6736(13)60843-0

7. Aboud FE, Yousafzai AK. Global Health and Development in Early Childhood. Annu Rev Psychol. 2015;66: 433–457. doi:10.1146/annurev-psych-010814-015128

8. Prado EL, Arnold CD, Wessells KR, Stewart CP, Abbeddou S, Adu-Afarwuah S, et al. Small-quantity lipid-based nutrient supplements for children age 6-24 months: a systematic review and individual participant data meta-analysis of effects on developmental outcomes and effect modifiers. Am J Clin Nutr. 2021;114: 43S–67S. doi:10.1093/ajcn/nqab277

9. Prado EL, Dewey KG. Nutrition and brain development in early life. Nutr Rev. 2014;72: 267–284. doi:10.1111/nure.12102

10. Pinkerton R, Oriá RB, Lima AAM, Rogawski ET, Oriá MOB, Patrick PD, et al. Early Childhood Diarrhea Predicts Cognitive Delays in Later Childhood Independently of Malnutrition. Am J Trop Med Hyg. 2016;95: 1004–1010. doi:10.4269/ajtmh.16-0150

11. Dearden KA, Brennan AT, Behrman JR, Schott W, Crookston BT, Humphries DL, et al. Does household access to improved water and sanitation in infancy and childhood predict better vocabulary test performance in Ethiopian, Indian, Peruvian and Vietnamese cohort studies? BMJ Open. 2017;7: e013201. doi:10.1136/bmjopen-2016-013201

12. Gladstone MJ, Chandna J, Kandawasvika G, Ntozini R, Majo FD, Tavengwa NV, et al. Independent and combined effects of improved water, sanitation, and hygiene (WASH) and improved complementary feeding on early neurodevelopment among children born to HIV-negative mothers in rural Zimbabwe: Substudy of a cluster-randomized trial. PLOS Med. 2019;16: e1002766. doi:10.1371/journal.pmed.1002766

13. Tofail F, Fernald LC, Das KK, Rahman M, Ahmed T, Jannat KK, et al. Effect of water quality, sanitation, hand washing, and nutritional interventions on child development in rural Bangladesh (WASH Benefits Bangladesh): a cluster-randomised controlled trial. Lancet Child Adolesc Health. 2018;2: 255–268. doi:10.1016/S2352-4642(18)30031-2

14. Stewart CP, Kariger P, Fernald L, Pickering AJ, Arnold CD, Arnold BF, et al. Effects of water quality, sanitation, handwashing, and nutritional interventions on child development in rural Kenya (WASH Benefits Kenya): a cluster-randomised controlled trial. Lancet Child Adolesc Health. 2018;2: 269–280. doi:10.1016/S2352-4642(18)30025-7

15. Jeong J, Pitchik HO, Fink G. Short-term, medium-term and long-term effects of early parenting interventions in low- and middle-income countries: a systematic review. BMJ Glob Health. 2021;6: e004067. doi:10.1136/bmjgh-2020-004067

16. Piper JD, Mazhanga C, Mwapaura M, Mapako G, Mapurisa I, Mashedze T, et al. School-age growth and development following infant feeding and/or water, sanitation, and hygiene interventions in rural Zimbabwe: long-term follow-up of a cluster-randomised trial. eClinicalMedicine. 2024;78. doi:10.1016/j.eclinm.2024.102946

17. Bowen A, Agboatwalla M, Luby S, Tobery T, Ayers T, Hoekstra RM. Association between intensive handwashing promotion and child development in Karachi, Pakistan: a cluster randomized controlled trial. Arch Pediatr Adolesc Med. 2012;166: 1037–1044. doi:10.1001/archpediatrics.2012.1181

18. Luby SP, Rahman M, Arnold BF, Unicomb L, Ashraf S, Winch PJ, et al. Effects of water quality, sanitation, handwashing, and nutritional interventions on diarrhoea and child growth in rural Bangladesh: a cluster randomised controlled trial. Lancet Glob Health. 2018;6: e302–e315. doi:10.1016/S2214-109X(17)30490-4

19. Arnold BF, Null C, Luby SP, Unicomb L, Stewart CP, Dewey KG, et al. Cluster-randomised controlled trials of individual and combined water, sanitation, hygiene and nutritional interventions in rural Bangladesh and Kenya: the WASH Benefits study design and rationale. BMJ Open. 2013;3: e003476. doi:10.1136/bmjopen-2013-003476

20. Wechsler D. Wechsler preschool and primary scale of intelligence—fourth edition. Psychol Corp San Antonio TX. 2012.

21. Ruan-Iu L, Pendergast LL, Rasheed M, Tofail F, Svensen E, Maphula A, et al. Assessing Early Childhood Fluid Reasoning in Low- and Middle-Income Nations: Validity of the Wechsler Preschool and Primary Scale of Intelligence Across Seven MAL-ED Sites. J Psychoeduc Assess. 2019; 0734282919850040. doi:10.1177/0734282919850040

22. Henderson SE, Sugden D, Barnett AL. Movement Assessment Battery for Children-2. APA PsycTests. 2007 [cited 27 Jun 2024]. doi:10.1037/t55281-000

23. Ali H, Hamadani J, Mehra S, Tofail F, Hasan MI, Shaikh S, et al. Effect of maternal antenatal and newborn supplementation with vitamin A on cognitive development of school-aged children in rural Bangladesh: a follow-up of a placebo-controlled, randomized trial. Am J Clin Nutr. 2017;106: 77–87. doi:10.3945/ajcn.116.134478

24. Goodman R. Psychometric Properties of the Strengths and Difficulties Questionnaire. J Am Acad Child Adolesc Psychiatry. 2001;40: 1337–1345. doi:10.1097/00004583-200111000-00015

25. Gustin K, Tofail F, Vahter M, Kippler M. Cadmium exposure and cognitive abilities and behavior at 10 years of age: A prospective cohort study. Environ Int. 2018;113: 259–268. doi:10.1016/j.envint.2018.02.020

26. Korkman M, Kirk U, Kemp S. NEPSY - Second Edition. APA PsycTests. 2007 [cited 27 Jun 2024]. doi:10.1037/t15125-000

27. Obradović J, Portilla XA, Boyce WT. Executive functioning and developmental neuroscience: Current progress and implications for early childhood education. Handb Early Child Educ. 2012; 324–351.

28. Wilkinson GS. Wide range achievement test 3: WRAT3. Wilmington, DE: Wide Range, Inc.; 1993. Available: https://search.library.wisc.edu/catalog/999714707202121

29. Hossain SJ, Tofail F, Mehrin SF, Hamadani JD. Six-Year Follow-up of Childhood Stimulation on Development of Children With and Without Anemia. Pediatrics. 2023;151: e2023060221E. doi:10.1542/peds.2023-060221E

30. Radloff LS. The CES-D Scale: A Self-Report Depression Scale for Research in the General Population. Appl Psychol Meas. 1977;1: 385–401. doi:10.1177/014662167700100306

31. Black MM, Baqui AH, Zaman K, Arifeen SE, Black RE. Maternal depressive symptoms and infant growth in rural Bangladesh. Am J Clin Nutr. 2009;89: 951S–957S. doi:10.3945/ajcn.2008.26692E

32. Bradley RH, Caldwell BM, Rock SL, Hamrick HM, Harris P. Home Observation for Measurement of the Environment: Development of a Home Inventory for use with families having children 6 to 10 years old. Contemp Educ Psychol. 1988;13: 58–71. doi:10.1016/0361-476X(88)90006-9

33. R Core Team. R: A language and environment for statistical computing. R Foundation for Statistical Computing, Vienna, Austria; 2022.

34. Kakwangire P, Muhoozi G, Ngari M, Matovu N, Westerberg AC, Iversen PO, et al. 8-Year Follow-up of a Maternal Education Trial in a Low-Resource Setting. Pediatrics. 2024; e2023063352. doi:10.1542/peds.2023-063352

35. Kakwangire P, Atukunda P, Ngari M, Westerberg AC, Iversen PO, Muhoozi G. Long-term effects on depressive symptoms among Ugandan mothers – Findings from a follow-up of a cluster-randomized education trial in a rural low-resource setting. J Affect Disord. 2024;351: 598–606. doi:10.1016/j.jad.2024.01.263

36. Yousafzai AK, Obradović J, Rasheed MA, Rizvi A, Portilla XA, Tirado-Strayer N, et al. Effects of responsive stimulation and nutrition interventions on children’s development and growth at age 4 years in a disadvantaged population in Pakistan: a longitudinal follow-up of a cluster-randomised factorial effectiveness trial. Lancet Glob Health. 2016;4: e548–558. doi:10.1016/S2214-109X(16)30100-0

37. Ocansey ME, Adu-Afarwuah S, Kumordzie SM, Okronipa H, Young RR, Tamakloe SM, et al. Prenatal and postnatal lipid-based nutrient supplementation and cognitive, social-emotional, and motor function in preschool-aged children in Ghana: a follow-up of a randomized controlled trial. Am J Clin Nutr. 2019;109: 322–334. doi:10.1093/ajcn/nqy303

38. Prado EL, Adu-Afarwuah S, Arnold CD, Adjetey E, Amponsah B, Bentil H, et al. Prenatal and postnatal small-quantity lipid-based nutrient supplements and children’s social–emotional difficulties at ages 9–11 y in Ghana: follow-up of a randomized controlled trial. Am J Clin Nutr. 2023;118: 433–442. doi:10.1016/j.ajcnut.2023.05.025

39. Prado EL, Sebayang SK, Apriatni M, Adawiyah SR, Hidayati N, Islamiyah A, et al. Maternal multiple micronutrient supplementation and other biomedical and socioenvironmental influences on children’s cognition at age 9–12 years in Indonesia: follow-up of the SUMMIT randomised trial. Lancet Glob Health. 2017;5: e217–e228. doi:10.1016/S2214-109X(16)30354-0

40. Grantham-McGregor SM, Walker SP, Chang SM, Powell CA. Effects of early childhood supplementation with and without stimulation on later development in stunted Jamaican children. Am J Clin Nutr. 1997;66: 247–253. doi:10.1093/ajcn/66.2.247

41. Cusick SE, Georgieff MK. The Role of Nutrition in Brain Development: The Golden Opportunity of the “First 1000 Days.” J Pediatr. 2016;175: 16–21. doi:10.1016/j.jpeds.2016.05.013

42. Niehaus MD, Moore SR, Patrick PD, Derr LL, Lorntz B, Lima AA, et al. Early childhood diarrhea is associated with diminished cognitive function 4 to 7 years later in children in a northeast Brazilian shantytown. Am J Trop Med Hyg. 2002;66: 590–593. doi:10.4269/ajtmh.2002.66.590

43. Petri WA, Miller M, Binder HJ, Levine MM, Dillingham R, Guerrant RL. Enteric infections, diarrhea, and their impact on function and development. J Clin Invest. 2008;118: 1277–1290. doi:10.1172/JCI34005

44. Rogawski McQuade ET, Platts-Mills JA, Gratz J, Zhang J, Moulton LH, Mutasa K, et al. Impact of Water Quality, Sanitation, Handwashing, and Nutritional Interventions on Enteric Infections in Rural Zimbabwe: The Sanitation Hygiene Infant Nutrition Efficacy (SHINE) Trial. J Infect Dis. 2020;221: 1379–1386. doi:10.1093/infdis/jiz179

45. Null C, Stewart CP, Pickering AJ, Dentz HN, Arnold BF, Arnold CD, et al. Effects of water quality, sanitation, handwashing, and nutritional interventions on diarrhoea and child growth in rural Kenya: a cluster-randomised controlled trial. Lancet Glob Health. 2018;6: e316–e329. doi:10.1016/S2214-109X(18)30005-6

46. Parvez SM, Azad R, Rahman M, Unicomb L, Ram PK, Naser AM, et al. Achieving optimal technology and behavioral uptake of single and combined interventions of water, sanitation hygiene and nutrition, in an efficacy trial (WASH benefits) in rural Bangladesh. Trials. 2018;19: 358. doi:10.1186/s13063-018-2710-8

47. Jeong J, Pitchik HO, Yousafzai AK. Stimulation Interventions and Parenting in Low- and Middle-Income Countries: A Meta-analysis. Pediatrics. 2018; e20173510. doi:10.1542/peds.2017-3510

