## Supplementary materials for "Effects of an early water, sanitation, hygiene, and nutritional intervention on child development at school age: a 7-year follow-up of a cluster-randomized trial in rural Bangladesh"

**Supplementary Materials for manuscript titled:** Effects of an early water, sanitation, hygiene, and nutritional intervention on child development at school age: a 7-year follow-up of a cluster-randomized trial in rural Bangladesh

*Fahmida Tofail*, Helen O. Pitchik*, Mahfuza Islam, Rizwana Khan, Abul K. Shoab, Fahmida Akter, Shirina Aktar, Tarique M.N. Huda, Mahbubur Rahman, Peter J. Winch, Stephen P. Luby, Lia C.H. Fernald*

### Table S1. WASH-B intervention components by intervention arm

| | Water | Sanitation | Handwashing | Nutrition | Water, Sanitation, Handwashing | Water, Sanitation, Handwashing, and nutrition |
|---|---|---|---|---|---|---|
| **Technology and supplies provided** | Insulated storage container for drinking water; Aquatabs (Medentech, Ireland) | Sani-scoop; potty; double-pit pour flush improved latrine | Handwashing station; storage bottle for soapy water; laundry detergent sachets for preparation of soapy water | LNS (Nutriset, France); storage container for LNS | Same as individual water, sanitation, and handwashing interventions | Same as individual water, sanitation, handwashing, and nutrition interventions |
| **Key behavioural recommendations delivered by promoters** | Targeted children drink treated, safely stored water | Family use double pit latrines; potty train children; safely dispose of faeces into latrine or pit | Family wash hands with soap after defecation and during food preparation | Exclusive breastfeeding up to 180 days; introduce diverse complementary food at 6 months; feed LNS from 6–24 months | Same as individual water, sanitation, and handwashing interventions | Same as individual water, sanitation, handwashing, and nutrition interventions |
| **Population targeted** | Children younger than 5 years living in index households | Whole compound for latrines; index households for potty training and safe faeces disposal | Residents of index households | Index children (targeted through mother) | Same as individual water, sanitation, and handwashing interventions | Same as individual water, sanitation, handwashing, and nutrition interventions |
| **Emphasis during visits after refresher training** | Safe storage of water, children drink only treated and safely stored water | Latrine cleanliness; maintenance; pit switching | Handwashing before food preparation | Dietary diversity during complementary feeding; provide LNS even if child is unwell | Same as individual water, sanitation, and handwashing interventions | Same as individual water, sanitation, handwashing, and nutrition interventions |

LNS=lipid-based nutrient supplement. †Promoter visits were intended to teach participants how to use technologies and how to use and restock products; arrange for social support; communicate benefits of use and practice and changes in social norms; congratulate and encourage; problem-solve as needed; and inspire. Techniques used included counselling via flipcharts and cue cards, onsite demonstrations of technologies and products, video dramas, storytelling, games, and songs. Promoter's guides detailed the visit objective, target audience, and the specific steps and materials to be used.

Table reproduced with minor adaptations from Luby et al 2018 (10.1016/S2214-109X(17)30490-4), with permission.

**Table S2. Comparing baseline characteristics for assessed vs. lost participants at the 5-year follow-up**

| No. of compounds | Assessed (N = 3835) | LTFU[1] (N = 1755) |
|---|---|---|
| **Maternal** | | |
| Years of education | 5.8 (3.4) | 5.8 (3.5) |
| **Paternal** | | |
| Years of education | 4.8 (4.0) | 4.9 (4.1) |
| Works in agriculture | 1262 (33%) | 493 (28%) |
| **Household** | | |
| Number of persons | 4.7 (2.1) | 4.6 (2.3) |
| Has electricity | 2332 (61%) | 962 (55%) |
| Has a cement floor | 412 (11%) | 177 (10%) |
| Acres of agricultural land owned | 0.2 (0.3) | 0.1 (0.2) |
| **Drinking Water** | | |
| Tubewell primary water source | 2856 (74%) | 1281 (73%) |
| Stored water observed at home | 1808 (47%) | 856 (49%) |
| **Sanitation** | | |
| Daily defecation in the open | | |
| Adult men | 272 (7%) | 146 (8%) |
| Adult women | 161 (4%) | 76 (4%) |
| Children: 8-<15 years | 162 (10%) | 69 (10%) |
| Children: 3-<8 years | 775 (38%) | 313 (38%) |
| Children: 0-<3 years | 665 (84%) | 309 (83%) |
| Latrine | | |
| Owned | 2046 (53%) | 950 (54%) |
| Concrete slab | 3466 (94%) | 1549 (93%) |
| Functional water seal | 964 (30%) | 420 (29%) |
| Visible stool on slab or floor | 1791 (50%) | 791 (48%) |
| Owned a potty | 157 (4%) | 88 (5%) |
| Human faeces observed in the | | |
| House | 328 (9%) | 138 (8%) |
| Child's play area | 44 (1%) | 20 (1%) |
| **Handwashing** | | |
| Within 6 steps of latrine | | |
| Has water | 425 (12%) | 184 (11%) |
| Has soap | 225 (6%) | 109 (7%) |
| Within 6 steps of kitchen | | |
| Has water | 297 (9%) | 149 (9%) |
| Has soap | 91 (3%) | 41 (3%) |

[1] Includes 34 participants who were followed-up and consented to participate, but were disabled and so did not complete the child development assessment

### Table S3. WPPSI results

| FSIQ z score | n | mean | unadj v C | adj v C | adj v N | adj v WSH |
|---|---|---|---|---|---|---|
| **Control** | 873 | 0.00 (1.00) | Ref | Ref | - | - |
| **Water** | 495 | 0.02 (1.03) | 0.00 (-0.10, 0.10) | 0.02 (-0.08, 0.13) | - | - |
| **Sanitation** | 484 | 0.16 (0.98) | 0.18 (0.05, 0.30) | 0.11 (-0.01, 0.22) | - | - |
| **Handwashing** | 492 | -0.02 (1.05) | -0.01 (-0.15, 0.13) | -0.02 (-0.15, 0.10) | - | - |
| **WSH** | 496 | 0.08 (0.98) | 0.06 (-0.06, 0.19) | 0.06 (-0.05, 0.17) | - | Ref |
| **Nutrition** | 494 | 0.05 (1.00) | 0.04 (-0.07, 0.15) | 0.07 (-0.03, 0.16) | Ref | - |
| **Nutrition+WSH** | 499 | 0.11 (1.00) | 0.11 (0.00, 0.21) | 0.12 (0.02, 0.23) | 0.05 (-0.09, 0.18) | 0.04 (-0.09, 0.17) |
| **CPI z score** | **n** | **mean** | **unadj v C** | **adj v C** | **adj v N** | **adj v WSH** |
| **Control** | 873 | 0.00 (1.00) | Ref | Ref | - | - |
| **Water** | 495 | -0.03 (1.02) | -0.03 (-0.14, 0.08) | -0.02 (-0.13, 0.09) | - | - |
| **Sanitation** | 484 | 0.07 (0.99) | 0.08 (-0.05, 0.21) | 0.03 (-0.09, 0.16) | - | - |
| **Handwashing** | 492 | -0.09 (1.05) | -0.08 (-0.22, 0.05) | -0.08 (-0.21, 0.05) | - | - |
| **WSH** | 496 | -0.02 (0.96) | -0.03 (-0.16, 0.09) | -0.04 (-0.14, 0.07) | - | Ref |
| **Nutrition** | 494 | -0.01 (0.99) | -0.01 (-0.12, 0.10) | 0.01 (-0.09, 0.11) | Ref | - |
| **Nutrition+WSH** | 499 | 0.08 (1.03) | 0.06 (-0.06, 0.17) | 0.09 (-0.02, 0.21) | 0.08 (-0.06, 0.22) | 0.11 (-0.02, 0.24) |
| **VCI z score** | **n** | **mean** | **unadj v C** | **adj v C** | **adj v N** | **adj v WSH** |
| **Control** | 873 | 0.00 (1.00) | Ref | Ref | - | - |
| **Water** | 495 | 0.08 (1.00) | 0.07 (-0.03, 0.17) | 0.07 (-0.03, 0.18) | - | - |
| **Sanitation** | 484 | 0.12 (1.01) | 0.14 (0.00, 0.28) | 0.07 (-0.05, 0.20) | - | - |
| **Handwashing** | 492 | 0.03 (1.06) | 0.04 (-0.10, 0.18) | 0.01 (-0.11, 0.14) | - | - |
| **WSH** | 496 | 0.05 (0.97) | 0.05 (-0.08, 0.18) | 0.03 (-0.09, 0.15) | - | Ref |
| **Nutrition** | 494 | 0.08 (1.01) | 0.06 (-0.05, 0.18) | 0.10 (0.00, 0.20) | Ref | - |
| **Nutrition+WSH** | 499 | 0.09 (0.98) | 0.10 (-0.01, 0.22) | 0.10 (-0.01, 0.22) | -0.01 (-0.15, 0.12) | 0.05 (-0.09, 0.18) |
| **FRI z score** | **n** | **mean** | **unadj v C** | **adj v C** | **adj v N** | **adj v WSH** |
| **Control** | 873 | 0.00 (1.00) | Ref | Ref | - | - |
| **Water** | 495 | 0.07 (1.01) | 0.06 (-0.05, 0.17) | 0.08 (-0.04, 0.19) | - | - |
| **Sanitation** | 484 | 0.16 (0.99) | 0.18 (0.07, 0.30) | 0.12 (0.00, 0.24) | - | - |
| **Handwashing** | 492 | -0.02 (1.02) | -0.02 (-0.16, 0.12) | -0.03 (-0.16, 0.10) | - | - |
| **WSH** | 496 | 0.10 (0.96) | 0.08 (-0.04, 0.20) | 0.10 (-0.01, 0.21) | - | Ref |
| **Nutrition** | 494 | 0.11 (1.05) | 0.11 (0.00, 0.23) | 0.12 (0.01, 0.23) | Ref | - |
| **Nutrition+WSH** | 499 | 0.15 (0.98) | 0.16 (0.04, 0.27) | 0.15 (0.03, 0.27) | 0.02 (-0.10, 0.14) | 0.06 (-0.05, 0.17) |
| **GAI z score** | **n** | **mean** | **unadj v C** | **adj v C** | **adj v N** | **adj v WSH** |
| **Control** | 873 | 0.00 (1.00) | Ref | Ref | - | - |
| **Water** | 495 | 0.05 (1.01) | 0.03 (-0.07, 0.13) | 0.05 (-0.06, 0.15) | - | - |
| **Sanitation** | 484 | 0.17 (0.99) | 0.19 (0.07, 0.31) | 0.12 (0.01, 0.23) | - | - |
| **Handwashing** | 492 | -0.01 (1.04) | -0.01 (-0.14, 0.13) | -0.02 (-0.14, 0.10) | - | - |
| **WSH** | 496 | 0.11 (0.97) | 0.09 (-0.04, 0.21) | 0.08 (-0.03, 0.20) | - | Ref |
| **Nutrition** | 494 | 0.09 (0.99) | 0.07 (-0.04, 0.19) | 0.11 (0.01, 0.20) | Ref | - |
| **Nutrition+WSH** | 499 | 0.11 (0.97) | 0.12 (0.02, 0.22) | 0.12 (0.03, 0.22) | 0.01 (-0.13, 0.14) | 0.02 (-0.11, 0.14) |
| **NVI z score** | **n** | **mean** | **unadj v C** | **adj v C** | **adj v N** | **adj v WSH** |
| **Control** | 873 | 0.00 (1.00) | Ref | Ref | - | - |
| **Water** | 495 | 0.01 (1.02) | -0.01 (-0.11, 0.09) | 0.02 (-0.09, 0.12) | - | - |
| **Sanitation** | 484 | 0.16 (0.97) | 0.18 (0.07, 0.30) | 0.11 (0.00, 0.22) | - | - |
| **Handwashing** | 492 | -0.04 (1.04) | -0.03 (-0.17, 0.11) | -0.04 (-0.16, 0.08) | - | - |
| **WSH** | 496 | 0.09 (0.98) | 0.07 (-0.06, 0.19) | 0.07 (-0.04, 0.18) | - | Ref |
| **Nutrition** | 494 | 0.03 (1.02) | 0.03 (-0.08, 0.14) | 0.05 (-0.05, 0.14) | Ref | - |
| **Nutrition+WSH** | 499 | 0.12 (1.01) | 0.11 (0.00, 0.22) | 0.13 (0.03, 0.24) | 0.07 (-0.06, 0.21) | 0.05 (-0.09, 0.18) |
| **WMI z score** | **n** | **mean** | **unadj v C** | **adj v C** | **adj v N** | **adj v WSH** |
| **Control** | 873 | 0.00 (1.00) | Ref | Ref | - | - |
| **Water** | 495 | 0.01 (1.03) | 0.02 (-0.10, 0.14) | 0.02 (-0.10, 0.13) | - | - |
| **Sanitation** | 484 | 0.09 (1.03) | 0.10 (-0.03, 0.24) | 0.05 (-0.08, 0.18) | - | - |
| **Handwashing** | 492 | -0.09 (1.06) | -0.12 (-0.26, 0.02) | -0.09 (-0.22, 0.04) | - | - |
| **WSH** | 496 | 0.04 (0.97) | 0.02 (-0.11, 0.15) | 0.04 (-0.08, 0.15) | - | Ref |
| **Nutrition** | 494 | 0.05 (0.99) | 0.04 (-0.08, 0.16) | 0.05 (-0.06, 0.17) | Ref | - |
| **Nutrition+WSH** | 499 | 0.05 (1.02) | 0.01 (-0.11, 0.13) | 0.06 (-0.06, 0.17) | 0.00 (-0.16, 0.16) | 0.01 (-0.13, 0.14) |

All outcomes are presented as internally standardized z-scores

FSIQ: Wechsler preschool and primary scales of intelligence Full Scale IQ; CPI: Cognitive Processing Index; VCI: Verbal Comprehension Index; FRI: Fluid reasoning index; GAI: General abilities index; NVI: Non-verbal index; WMI: Working memory index

### Table S4. Fine motor, Executive functioning, School achievement, SDQ results

| MABC MD | n | mean | unadj v C | adj v C | adj v N | adj v WSH |
|---|---|---|---|---|---|---|
| Control | 854 | 0.00 (1.00) | Ref | Ref | - | - |
| Water | 477 | -0.04 (0.95) | -0.10 (-0.27, 0.08) | -0.03 (-0.15, 0.09) | - | - |
| Sanitation | 469 | 0.05 (1.00) | 0.05 (-0.09, 0.19) | 0.04 (-0.08, 0.16) | - | - |
| Handwashing | 475 | 0.07 (1.06) | 0.12 (-0.08, 0.31) | 0.07 (-0.07, 0.21) | - | - |
| WSH | 478 | 0.06 (0.95) | 0.06 (-0.09, 0.21) | 0.09 (-0.03, 0.22) | - | Ref |
| Nutrition | 473 | 0.02 (0.92) | 0.06 (-0.11, 0.23) | 0.04 (-0.08, 0.16) | Ref | - |
| Nutrition+WSH | 488 | 0.05 (1.03) | 0.05 (-0.11, 0.22) | 0.06 (-0.06, 0.18) | 0.00 (-0.14, 0.14) | -0.03 (-0.17, 0.12) |
| **MABC MD1** | **n** | **mean** | **unadj v C** | **adj v C** | **adj v N** | **adj v WSH** |
| Control | 873 | 0.00 (1.00) | Ref | Ref | - | - |
| Water | 495 | -0.05 (1.01) | -0.05 (-0.17, 0.07) | -0.05 (-0.17, 0.07) | - | - |
| Sanitation | 484 | 0.00 (0.97) | 0.00 (-0.11, 0.11) | -0.01 (-0.12, 0.11) | - | - |
| Handwashing | 491 | 0.06 (0.87) | 0.05 (-0.07, 0.17) | 0.04 (-0.08, 0.17) | - | - |
| WSH | 496 | -0.04 (1.01) | -0.03 (-0.14, 0.09) | -0.05 (-0.17, 0.08) | - | Ref |
| Nutrition | 494 | -0.04 (1.13) | -0.04 (-0.17, 0.09) | -0.05 (-0.18, 0.08) | Ref | - |
| Nutrition+WSH | 499 | 0.04 (0.95) | 0.04 (-0.08, 0.16) | 0.04 (-0.08, 0.17) | 0.08 (-0.06, 0.22) | 0.11 (-0.02, 0.24) |
| **MABC MD2** | **n** | **mean** | **unadj v C** | **adj v C** | **adj v N** | **adj v WSH** |
| Control | 872 | 0.00 (1.00) | Ref | Ref | - | - |
| Water | 495 | -0.04 (1.15) | -0.02 (-0.17, 0.13) | -0.02 (-0.16, 0.13) | - | - |
| Sanitation | 484 | 0.04 (0.99) | 0.03 (-0.11, 0.17) | 0.05 (-0.08, 0.19) | - | - |
| Handwashing | 491 | 0.03 (1.29) | 0.06 (-0.10, 0.22) | 0.04 (-0.11, 0.18) | - | - |
| WSH | 495 | 0.06 (0.99) | 0.10 (-0.03, 0.23) | 0.10 (-0.02, 0.22) | - | Ref |
| Nutrition | 494 | -0.01 (1.02) | 0.00 (-0.12, 0.12) | 0.02 (-0.10, 0.14) | Ref | - |
| Nutrition+WSH | 499 | 0.09 (1.07) | 0.05 (-0.08, 0.19) | 0.09 (-0.02, 0.21) | 0.06 (-0.11, 0.22) | -0.02 (-0.18, 0.13) |
| **MABC MD3** | **n** | **mean** | **unadj v C** | **adj v C** | **adj v N** | **adj v WSH** |
| Control | 854 | 0.00 (1.00) | Ref | Ref | - | - |
| Water | 477 | -0.04 (1.05) | -0.06 (-0.22, 0.09) | -0.03 (-0.14, 0.09) | - | - |
| Sanitation | 469 | 0.02 (1.22) | 0.07 (-0.07, 0.21) | 0.02 (-0.11, 0.14) | - | - |
| Handwashing | 475 | 0.04 (1.06) | 0.12 (-0.05, 0.29) | 0.05 (-0.09, 0.18) | - | - |
| WSH | 479 | 0.09 (0.93) | 0.02 (-0.13, 0.18) | 0.11 (-0.01, 0.23) | - | Ref |
| Nutrition | 473 | 0.07 (0.91) | 0.11 (-0.04, 0.26) | 0.08 (-0.03, 0.20) | Ref | - |
| Nutrition+WSH | 488 | -0.03 (1.17) | 0.01 (-0.14, 0.17) | -0.02 (-0.15, 0.11) | -0.10 (-0.25, 0.05) | -0.11 (-0.26, 0.04) |
| **NEPSY FC** | **n** | **mean** | **unadj v C** | **adj v C** | **adj v N** | **adj v WSH** |
| Control | 873 | -0.01 (1.00) | Ref | Ref | - | - |
| Water | 495 | 0.04 (1.07) | 0.04 (-0.08, 0.17) | 0.05 (-0.08, 0.17) | - | - |
| Sanitation | 484 | 0.10 (1.08) | 0.12 (-0.01, 0.26) | 0.06 (-0.07, 0.19) | - | - |
| Handwashing | 492 | 0.02 (1.05) | 0.06 (-0.07, 0.19) | 0.04 (-0.09, 0.16) | - | - |
| WSH | 496 | -0.03 (0.98) | 0.01 (-0.11, 0.13) | -0.04 (-0.16, 0.07) | - | Ref |
| Nutrition | 494 | 0.07 (0.98) | 0.07 (-0.04, 0.18) | 0.08 (-0.03, 0.19) | Ref | - |
| Nutrition+WSH | 499 | 0.08 (0.99) | 0.11 (-0.01, 0.23) | 0.10 (-0.01, 0.21) | 0.03 (-0.11, 0.18) | 0.12 (-0.02, 0.25) |
| **FWS** | **n** | **mean** | **unadj v C** | **adj v C** | **adj v N** | **adj v WSH** |
| Control | 873 | 0.00 (1.00) | Ref | Ref | - | - |
| Water | 495 | 0.08 (1.09) | 0.08 (-0.04, 0.20) | 0.03 (-0.09, 0.15) | - | - |
| Sanitation | 484 | 0.10 (1.04) | 0.12 (0.01, 0.24) | 0.06 (-0.06, 0.18) | - | - |
| Handwashing | 492 | -0.02 (1.01) | -0.04 (-0.16, 0.07) | -0.02 (-0.13, 0.09) | - | - |
| WSH | 496 | 0.09 (1.05) | 0.07 (-0.06, 0.20) | 0.03 (-0.09, 0.15) | - | Ref |
| Nutrition | 494 | 0.05 (0.98) | 0.03 (-0.08, 0.14) | 0.04 (-0.08, 0.16) | Ref | - |
| Nutrition+WSH | 499 | 0.02 (1.04) | 0.02 (-0.08, 0.12) | 0.08 (-0.04, 0.20) | 0.03 (-0.11, 0.16) | 0.05 (-0.09, 0.19) |
| **Corsi blocks** | **n** | **mean** | **unadj v C** | **adj v C** | **adj v N** | **adj v WSH** |
| Control | 873 | 0.00 (1.00) | Ref | Ref | - | - |
| Water | 495 | -0.02 (1.05) | 0.00 (-0.11, 0.11) | 0.01 (-0.11, 0.13) | - | - |
| Sanitation | 484 | 0.09 (0.98) | 0.10 (-0.01, 0.21) | 0.09 (-0.02, 0.19) | - | - |
| Handwashing | 492 | 0.00 (1.01) | 0.02 (-0.10, 0.14) | 0.02 (-0.10, 0.15) | - | - |
| WSH | 496 | 0.01 (1.02) | 0.03 (-0.09, 0.15) | 0.01 (-0.09, 0.12) | - | Ref |
| Nutrition | 494 | 0.02 (0.95) | 0.02 (-0.08, 0.12) | 0.03 (-0.06, 0.12) | Ref | - |
| Nutrition+WSH | 499 | 0.03 (0.98) | 0.05 (-0.06, 0.15) | 0.04 (-0.06, 0.15) | 0.00 (-0.13, 0.13) | 0.04 (-0.09, 0.18) |
| **WRAT spelling** | **n** | **mean** | **unadj v C** | **adj v C** | **adj v N** | **adj v WSH** |
| Control | 873 | 0.00 (1.00) | Ref | Ref | - | - |
| Water | 495 | -0.01 (1.02) | -0.03 (-0.14, 0.09) | 0.00 (-0.11, 0.11) | - | - |

| | | | | | | |
|---|---|---|---|---|---|---|
| **Sanitation** | 484 | 0.06 (0.99) | 0.05 (-0.07, 0.18) | 0.01 (-0.10, 0.13) | - | - |
| **Handwashing** | 492 | -0.03 (1.04) | -0.04 (-0.18, 0.09) | -0.04 (-0.15, 0.08) | - | - |
| **WSH** | 496 | 0.02 (1.03) | -0.01 (-0.14, 0.12) | 0.00 (-0.11, 0.11) | - | Ref |
| **Nutrition** | 494 | 0.04 (1.04) | 0.04 (-0.09, 0.16) | 0.03 (-0.09, 0.15) | Ref | - |
| **Nutrition+WSH** | 499 | 0.03 (1.00) | 0.00 (-0.12, 0.12) | 0.03 (-0.08, 0.14) | -0.02 (-0.14, 0.11) | 0.03 (-0.10, 0.16) |
| **WRAT math** | **n** | **mean** | **unadj v C** | **adj v C** | **adj v N** | **adj v WSH** |
| **Control** | 873 | 0.00 (1.01) | Ref | Ref | - | - |
| **Water** | 495 | 0.02 (1.06) | -0.01 (-0.13, 0.10) | 0.03 (-0.09, 0.15) | - | - |
| **Sanitation** | 484 | 0.11 (1.03) | 0.09 (-0.03, 0.21) | 0.06 (-0.06, 0.18) | - | - |
| **Handwashing** | 492 | -0.01 (1.05) | -0.03 (-0.15, 0.09) | -0.02 (-0.13, 0.09) | - | - |
| **WSH** | 496 | 0.05 (1.02) | 0.05 (-0.08, 0.18) | 0.03 (-0.09, 0.15) | - | Ref |
| **Nutrition** | 494 | 0.05 (1.03) | 0.04 (-0.08, 0.17) | 0.04 (-0.08, 0.16) | Ref | - |
| **Nutrition+WSH** | 499 | 0.07 (1.03) | 0.05 (-0.08, 0.18) | 0.08 (-0.04, 0.20) | 0.03 (-0.11, 0.16) | 0.05 (-0.09, 0.19) |
| **WRAT read** | **n** | **mean** | **unadj v C** | **adj v C** | **adj v N** | **adj v WSH** |
| **Control** | 873 | 0.01 (1.01) | Ref | Ref | - | - |
| **Water** | 495 | -0.01 (1.01) | -0.02 (-0.13, 0.08) | 0.00 (-0.11, 0.11) | - | - |
| **Sanitation** | 484 | 0.02 (0.97) | 0.03 (-0.10, 0.15) | -0.02 (-0.14, 0.10) | - | - |
| **Handwashing** | 492 | -0.03 (1.04) | -0.05 (-0.18, 0.08) | -0.04 (-0.16, 0.08) | - | - |
| **WSH** | 496 | 0.03 (1.05) | -0.01 (-0.16, 0.13) | 0.01 (-0.11, 0.13) | - | Ref |
| **Nutrition** | 494 | 0.02 (1.01) | 0.02 (-0.11, 0.14) | 0.01 (-0.11, 0.13) | Ref | - |
| **Nutrition+WSH** | 499 | 0.02 (1.00) | 0.00 (-0.13, 0.13) | 0.02 (-0.10, 0.14) | -0.01 (-0.13, 0.12) | 0.02 (-0.12, 0.16) |
| **SDQ difficulty** | **n** | **mean** | **unadj v C** | **adj v C** | **adj v N** | **adj v WSH** |
| **Control** | 874 | 0.00 (1.00) | Ref | Ref | - | - |
| **Water** | 495 | -0.07 (0.98) | -0.08 (-0.23, 0.07) | -0.07 (-0.22, 0.09) | - | - |
| **Sanitation** | 484 | 0.05 (0.90) | 0.02 (-0.11, 0.16) | 0.04 (-0.10, 0.17) | - | - |
| **Handwashing** | 492 | -0.02 (0.92) | -0.03 (-0.15, 0.10) | -0.02 (-0.15, 0.11) | - | - |
| **WSH** | 495 | -0.06 (0.99) | -0.06 (-0.20, 0.08) | -0.07 (-0.22, 0.07) | - | Ref |
| **Nutrition** | 494 | -0.06 (0.89) | -0.06 (-0.22, 0.09) | -0.07 (-0.22, 0.08) | Ref | - |
| **Nutrition+WSH** | 500 | 0.02 (0.94) | 0.03 (-0.10, 0.17) | 0.02 (-0.12, 0.15) | 0.08 (-0.06, 0.22) | 0.09 (-0.06, 0.25) |
| **SDQ prosocial** | **n** | **mean** | **unadj v C** | **adj v C** | **adj v N** | **adj v WSH** |
| **Control** | 874 | 0.02 (0.99) | Ref | Ref | - | - |
| **Water** | 495 | 0.19 (0.98) | 0.18 (0.06, 0.31) | 0.19 (0.06, 0.32) | - | - |
| **Sanitation** | 484 | 0.13 (1.02) | 0.12 (-0.02, 0.25) | 0.10 (-0.04, 0.23) | - | - |
| **Handwashing** | 492 | 0.22 (0.91) | 0.20 (0.07, 0.33) | 0.21 (0.08, 0.34) | - | - |
| **WSH** | 495 | 0.30 (0.96) | 0.27 (0.13, 0.42) | 0.29 (0.15, 0.43) | - | Ref |
| **Nutrition** | 494 | 0.19 (0.98) | 0.17 (0.03, 0.31) | 0.21 (0.07, 0.35) | Ref | - |
| **Nutrition+WSH** | 500 | 0.33 (1.00) | 0.31 (0.15, 0.46) | 0.32 (0.16, 0.48) | 0.12 (-0.05, 0.29) | 0.01 (-0.15, 0.17) |

All outcomes are presented as internally standardized z-scores.
MABC: MD: Movement assessment battery for children manual dexterity scores; MD: manual dexterity overall score; MD1: first measure of manual dexterity – posting coins or placing pegs (depending on child age); MD2: second measure of manual dexterity – Threading lace; MD3: third measure of manual dexterity – Drawing trails; SDQ: Strengths and difficulties questionnaire; WRAT: Wide Range Achievement Test

### Table S5. Secondary outcome results

| CES-D z-score | n | mean | unadj v C | adj v C | adj v N | adj v WSH |
|---|---|---|---|---|---|---|
| **Control** | 865 | 0.00 (1.00) | Ref | Ref | - | - |
| **Water** | 495 | 0.04 (1.06) | 0.00 (-0.12, 0.12) | 0.05 (-0.06, 0.16) | - | - |
| **Sanitation** | 483 | -0.06 (1.05) | -0.06 (-0.19, 0.06) | -0.05 (-0.17, 0.06) | - | - |
| **Handwashing** | 486 | -0.15 (0.98) | -0.19 (-0.31, -0.07) | -0.14 (-0.24, -0.03) | - | - |
| **WSH** | 496 | -0.04 (1.04) | -0.02 (-0.16, 0.11) | -0.03 (-0.16, 0.10) | - | Ref |
| **Nutrition** | 491 | -0.22 (0.93) | -0.22 (-0.33, -0.11) | -0.21 (-0.31, -0.11) | Ref | - |
| **Nutrition+WSH** | 496 | -0.14 (1.02) | -0.13 (-0.27, 0.01) | -0.15 (-0.27, -0.02) | 0.11 (-0.02, 0.24) | -0.09 (-0.24, 0.06) |
| **HOME z-score** | **n** | **mean** | **unadj v C** | **adj v C** | **adj v N** | **adj v WSH** |
| **Control** | 874 | 0.01 (0.99) | Ref | Ref | - | - |
| **Water** | 495 | 0.20 (0.99) | 0.20 (0.04, 0.37) | 0.19 (0.03, 0.35) | - | - |
| **Sanitation** | 484 | 0.23 (0.99) | 0.24 (0.08, 0.40) | 0.17 (0.00, 0.35) | - | - |
| **Handwashing** | 492 | 0.24 (0.92) | 0.26 (0.10, 0.41) | 0.26 (0.11, 0.42) | - | - |
| **WSH** | 496 | 0.39 (0.96) | 0.35 (0.20, 0.51) | 0.31 (0.15, 0.48) | - | Ref |
| **Nutrition** | 494 | 0.24 (1.01) | 0.26 (0.09, 0.43) | 0.24 (0.08, 0.41) | Ref | - |
| **Nutrition+WSH** | 500 | 0.41 (0.96) | 0.43 (0.27, 0.59) | 0.41 (0.24, 0.57) | 0.17 (-0.03, 0.37) | 0.05 (-0.15, 0.24) |

All outcomes are presented as internally standardized z-scores
CES-D: Center for epidemiologic studies 20-question depression measure; HOME: Middle childhood Home Observation Measurement of the Environment

**Figure S1. WPPSI index scores**

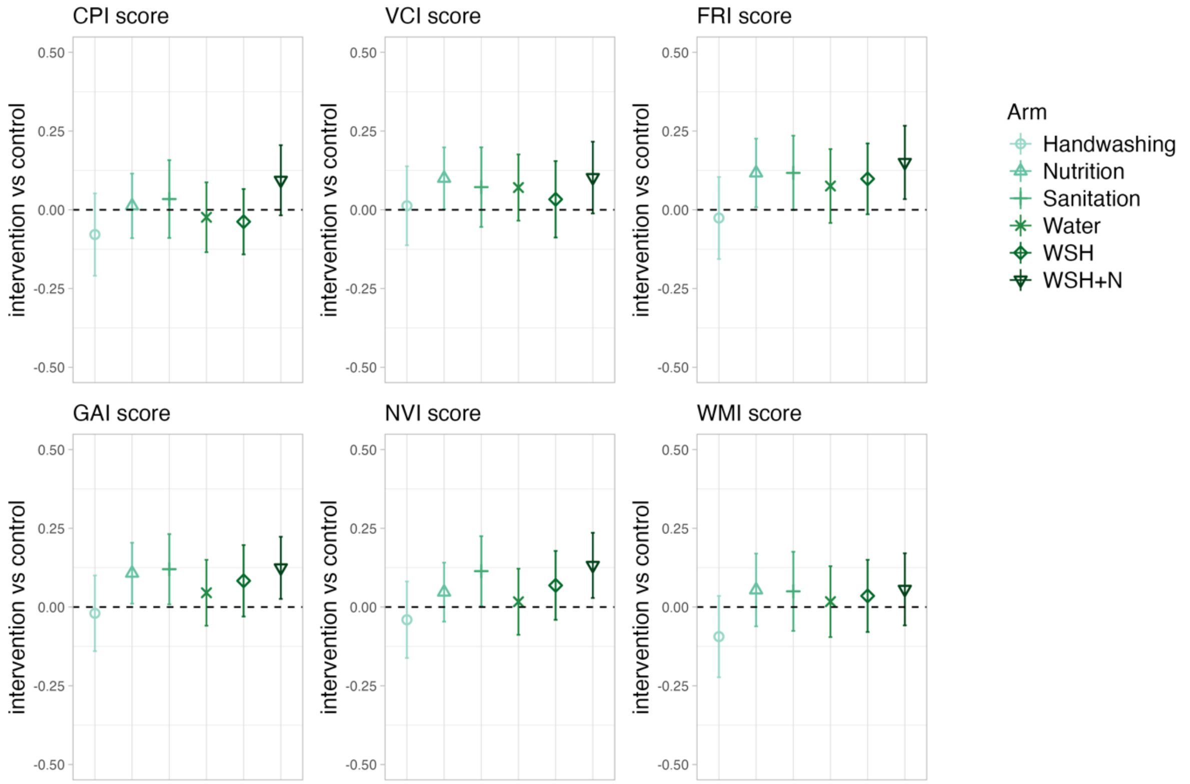


All outcomes are presented as internally standardized z-scores
CPI: Cognitive Processing Index; VCI: Verbal Comprehension Index; FRI: Fluid reasoning index; GAI: General abilities index; NVI: Non-verbal index; WMI: Working memory index

Note: for WPPSI I did not do the processing speed index as it is not a formal WPPSI calculation. (it is part of the statistical analysis plan though)

**Figure S2. Expanded narrative memory results**

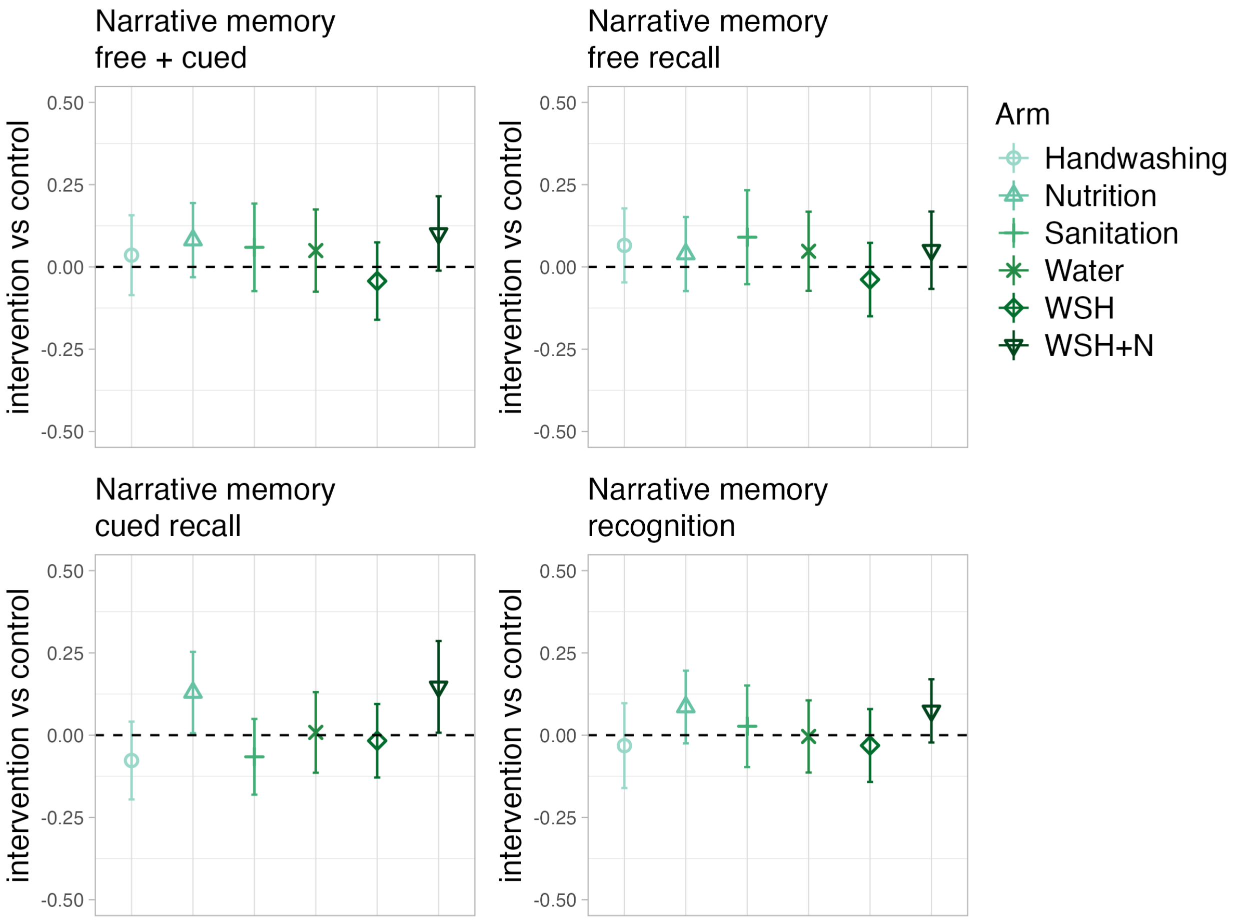

### Figure S3. Subgroup analysis for SDQ prosocial outcome

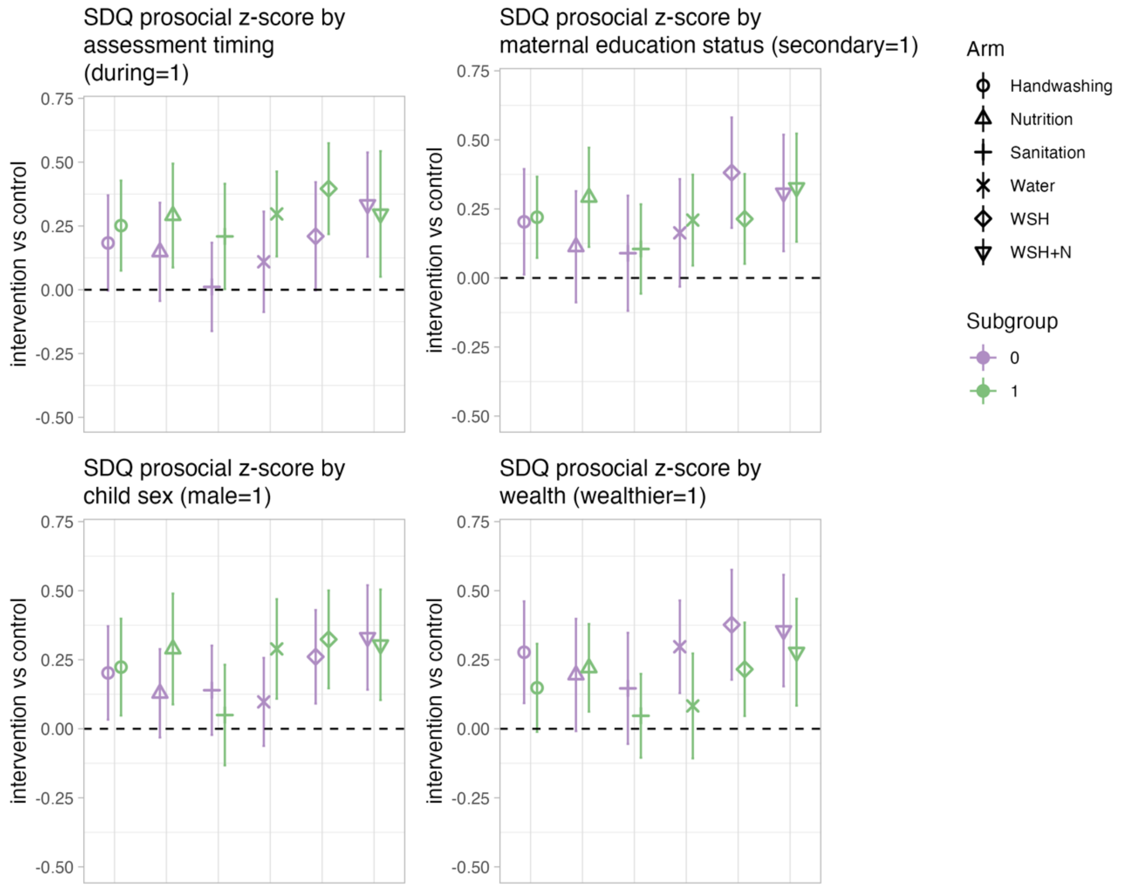

### Figure S4. Subgroup analysis for SDQ difficulties

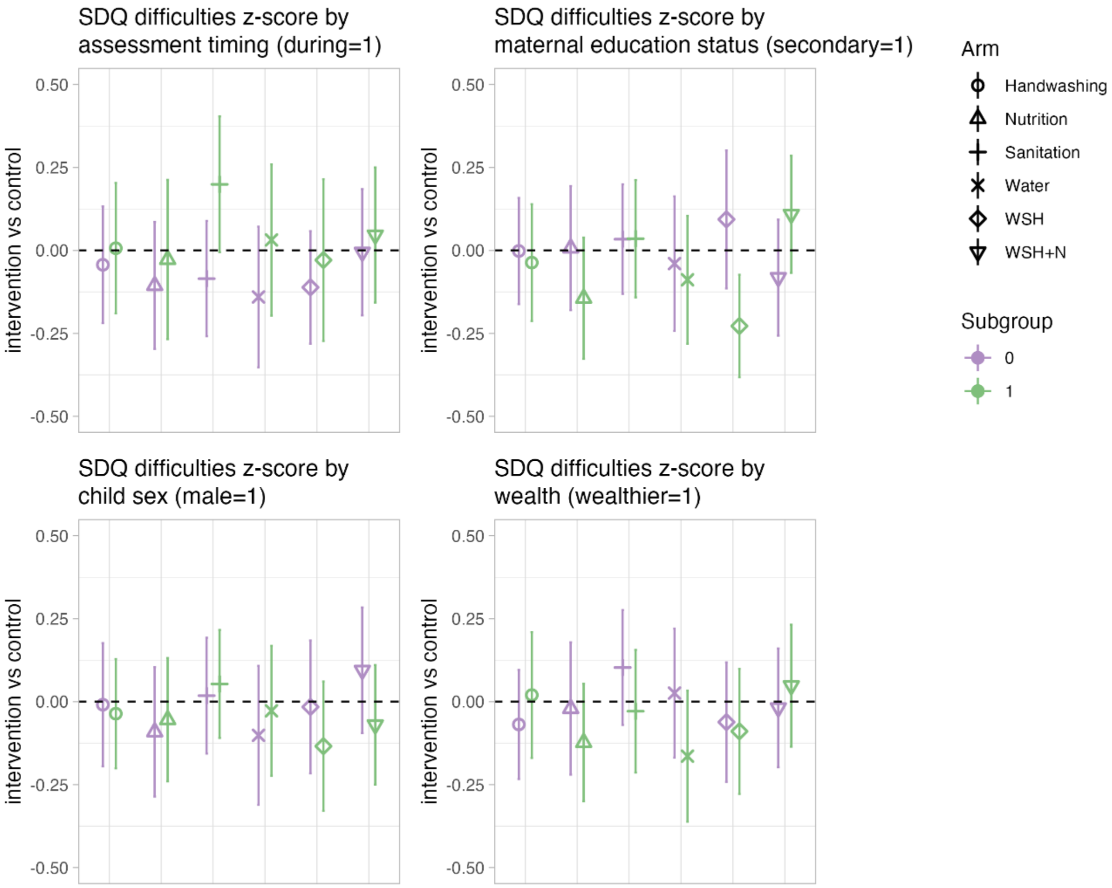

**Figure S5. Subgroup analysis for Fine motor outcome**

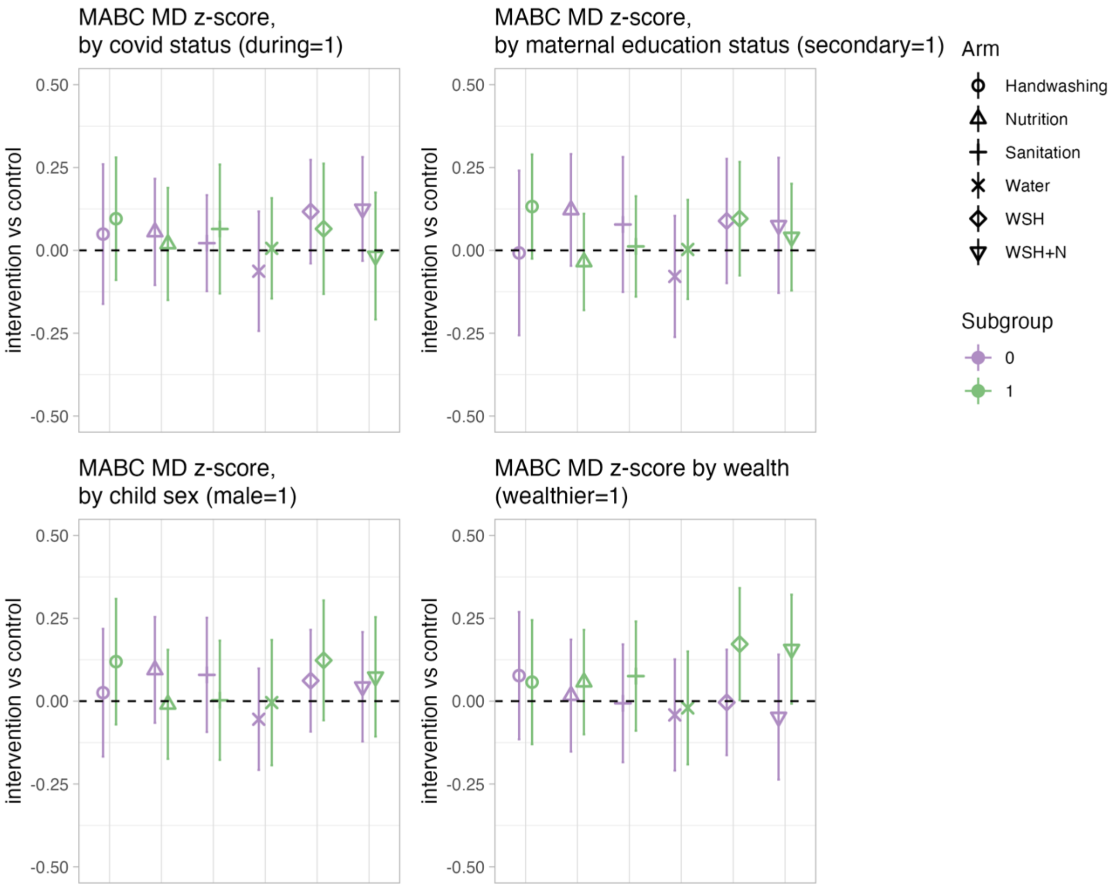

**Figure S6. Subgroup analysis for Math achievement**

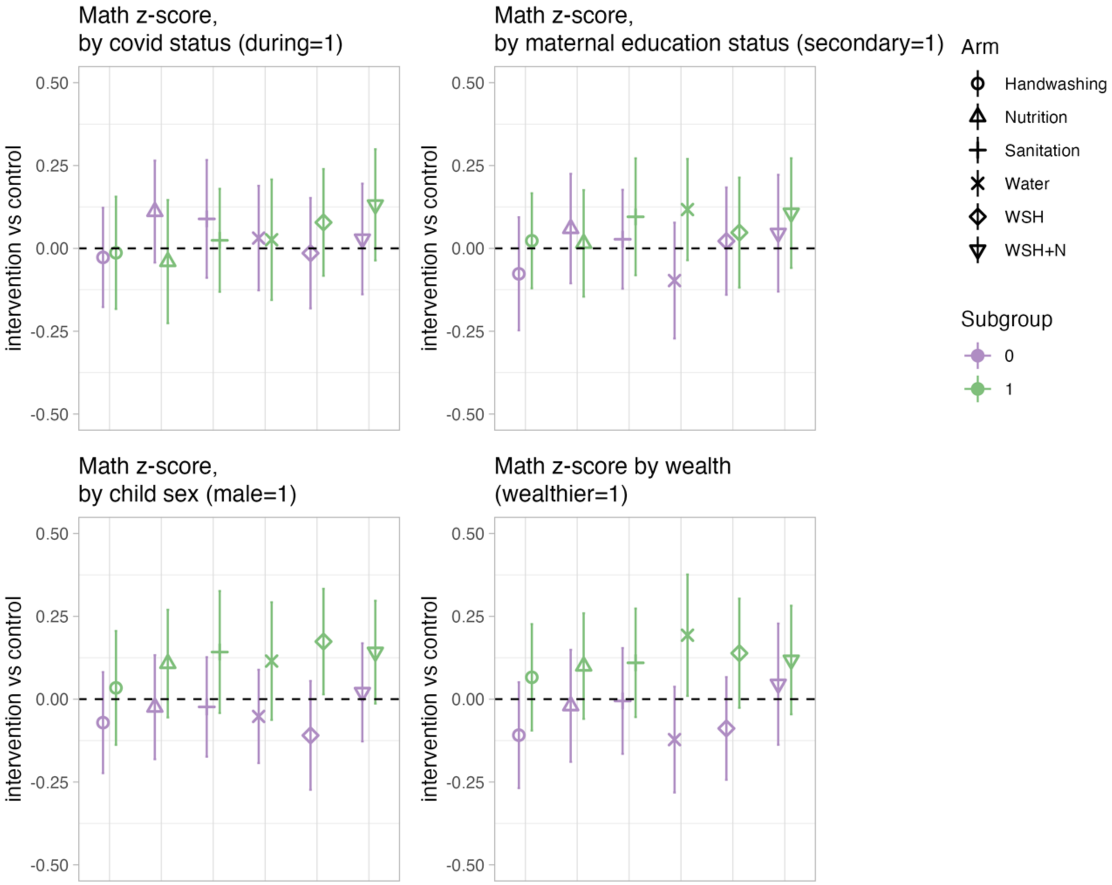

**Figure S7. Subgroup analysis for Reading achievement**

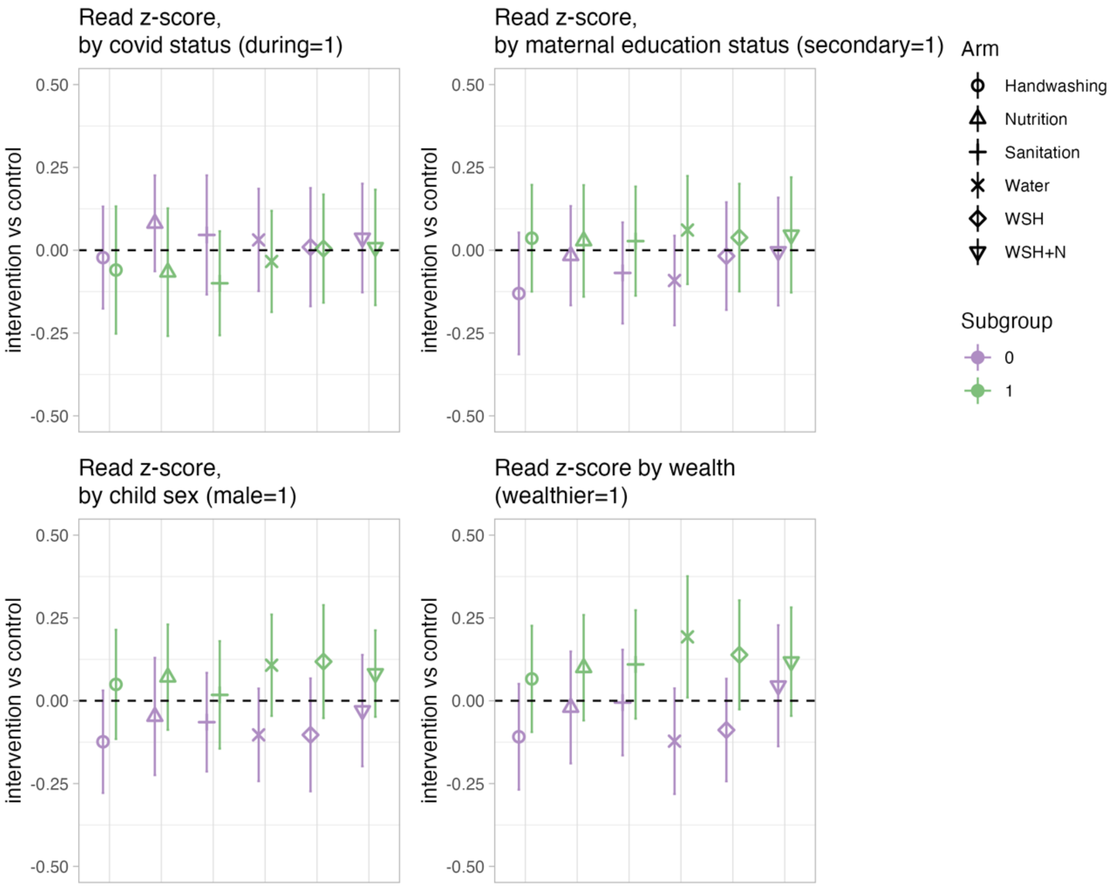

**Figure S8. Subgroup analysis for Spelling achievement**

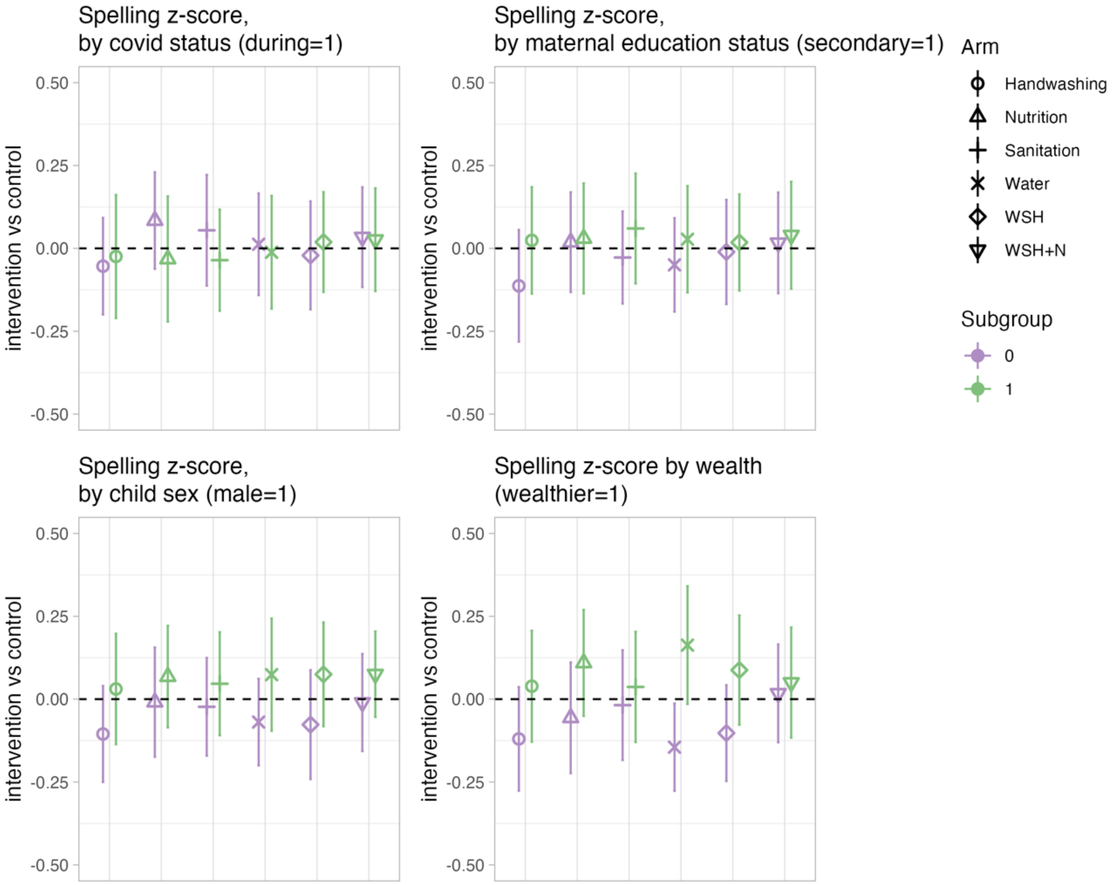

**Figure S9. Subgroup for Narrative memory**

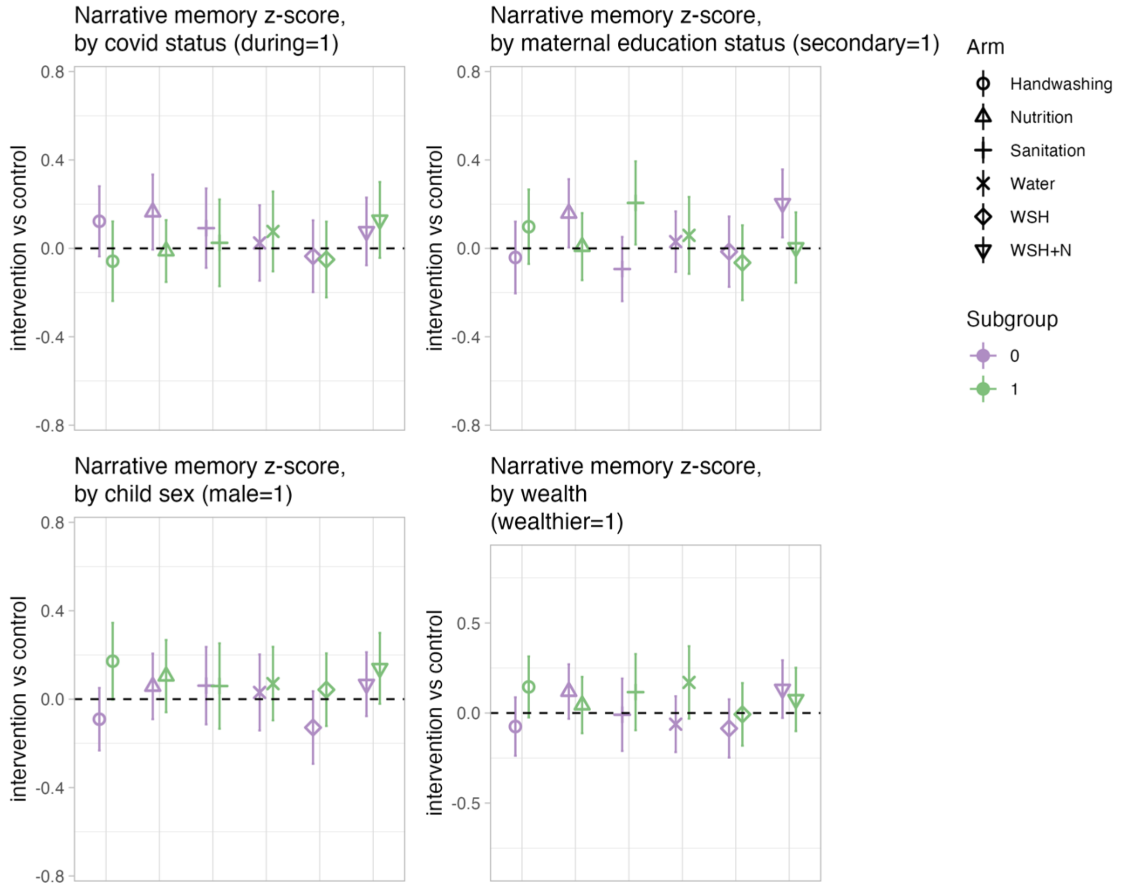

### Figure S10. Subgroup for Corsi blocks

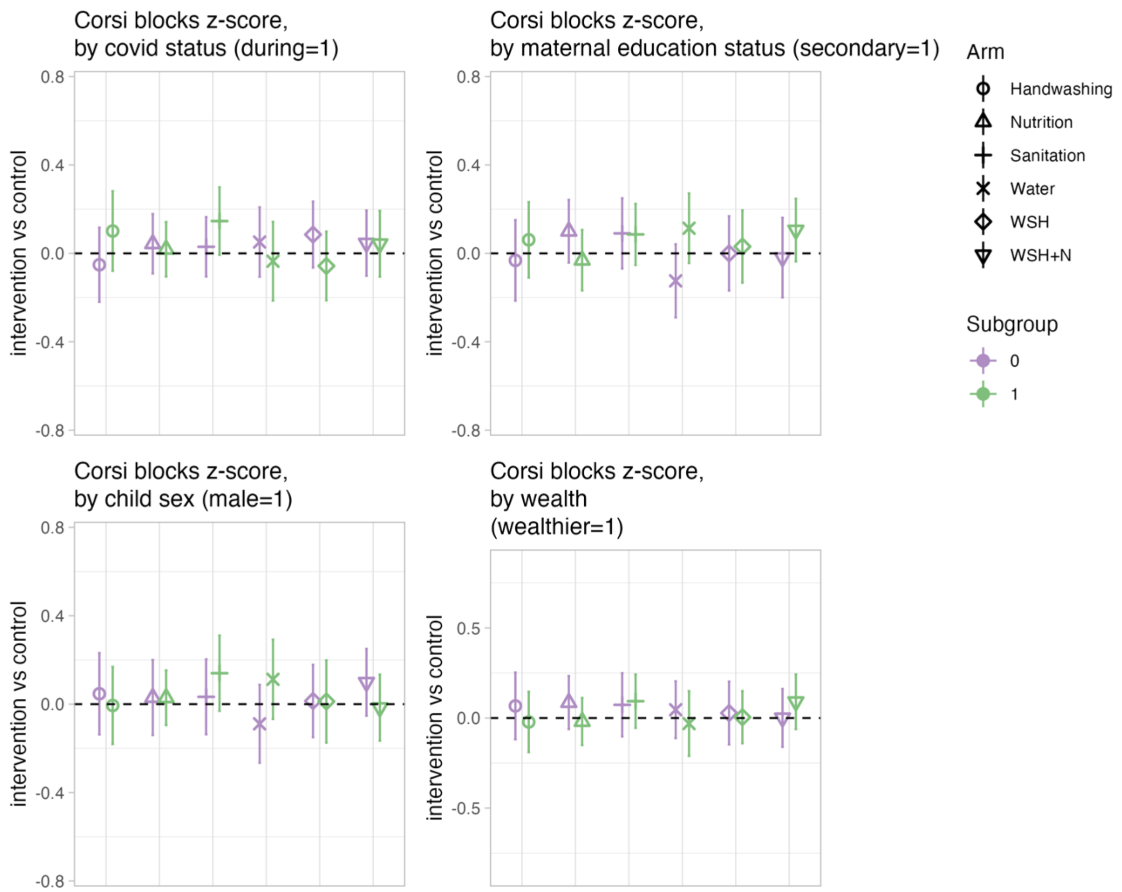

### Figure S11. Subgroup for Forward word span

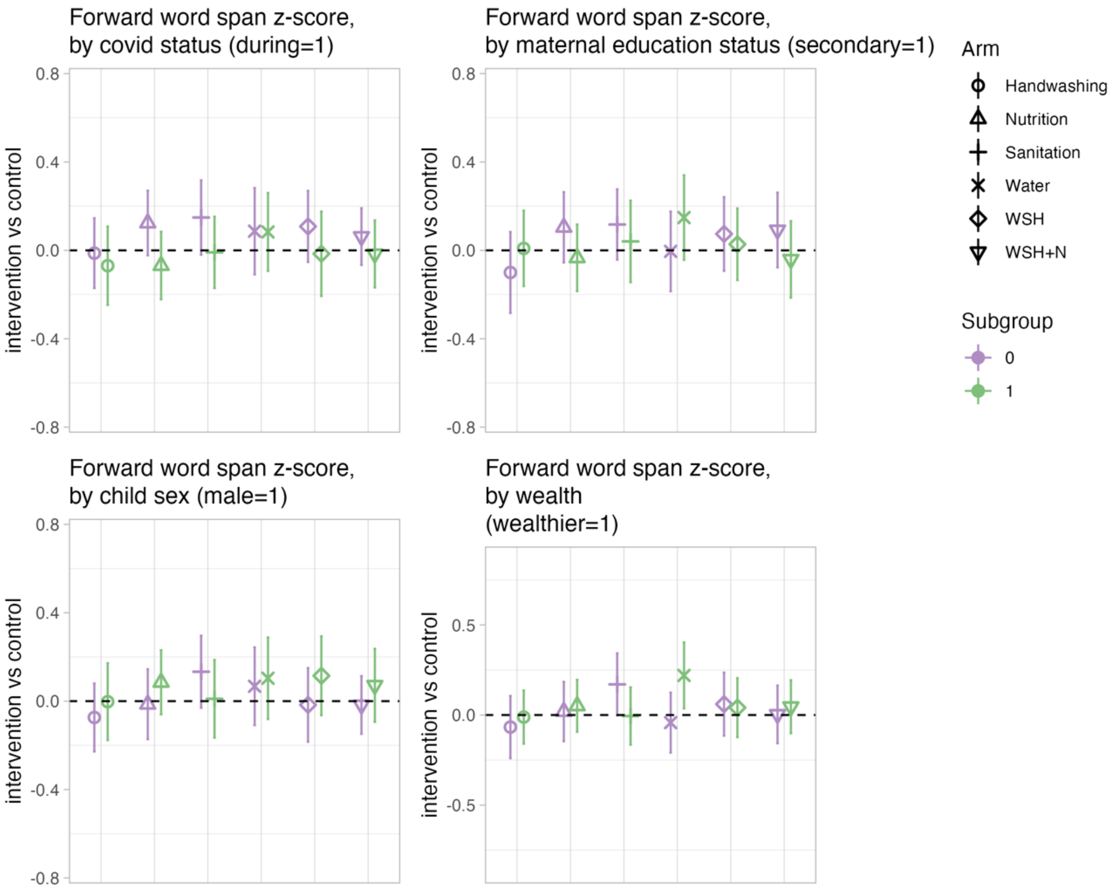

**Figure S12. Subgroup analysis for HOME**

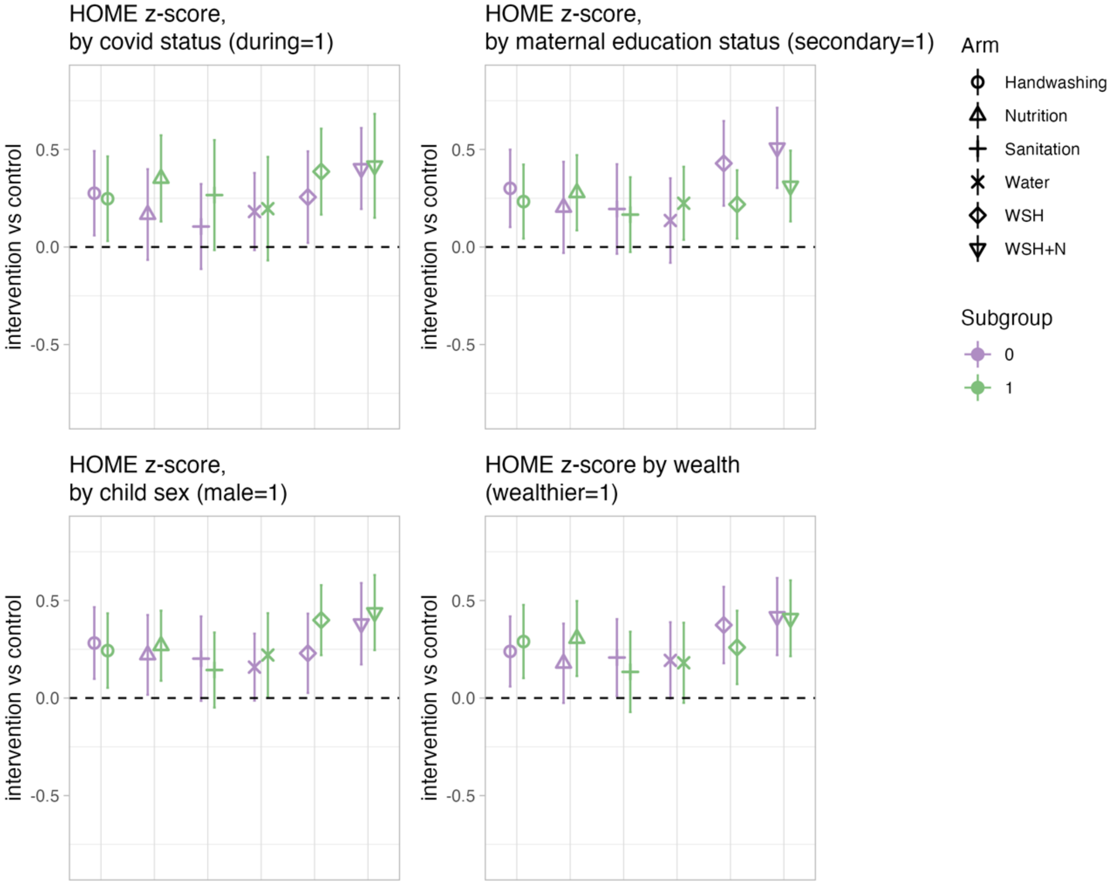

**Figure S13. Subgroup analysis for Maternal Depressive symptoms**

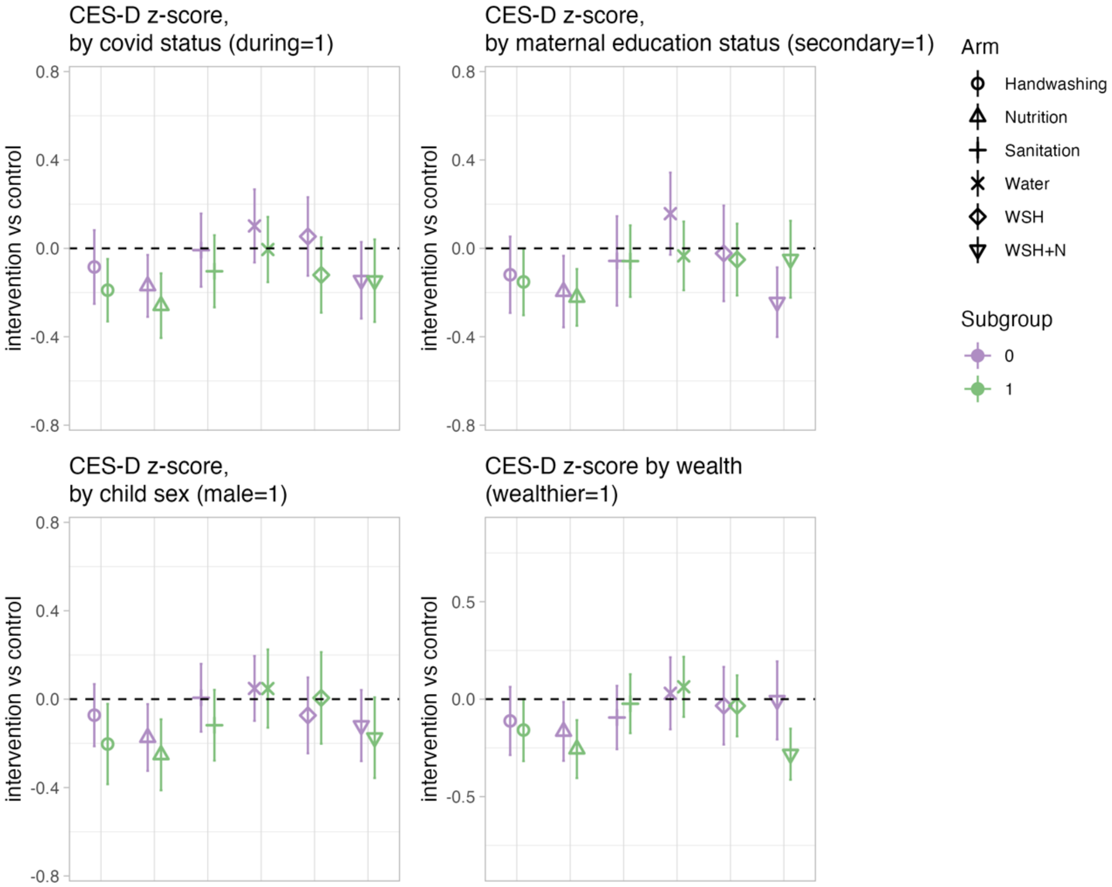
